# Endocannabinoid system genes and dementia: a systematic Mendelian randomization study

**DOI:** 10.64898/2026.09.08.26362514

**Authors:** Sara Javidnia, Ville Karhunen, Benjamin Woolf, Stephen Burgess, Petra Proitsi, Latha Velayudhan, Sagnik Bhattacharyya

**Affiliations:** Department of Psychosis Studies, Institute of Psychiatry, Psychology and Neuroscience, King’s College London, London, UK; MRC Biostatistics Unit, University of Cambridge, Cambridge, UK; Department of Public Health and Primary Care, School of Clinical Medicine, University of Cambridge, Cambridge, UK; Centre for Preventive Neurology, Wolfson Institute of Population Health, Queen Mary University of London, London, UK; Department of Basic and Clinical Neuroscience, King’s College London, London, UK; Centre for Healthy Brain Ageing, Department of Psychological Medicine, Institute of Psychiatry, Psychology and Neuroscience, King’s College London, UK

**Keywords:** Dementia, endocannabinoid system, Mendelian randomization, functional variants, tissue-specific *cis*-eQTL

## Abstract

**INTRODUCTION:** Dementia remains a major global health challenge, and human evidence linking endocannabinoid system (ECS) genetic variation to dementia is limited.

**METHODS:** We investigated seven ECS genes (*CNR1*, *CNR2*, *FAAH*, *DAGLA*, *DAGLB*, *NAPEPLD*, *MGLL*) in Alzheimer’s disease (AD), vascular dementia (VasD), and all-cause dementia. Functional variants were evaluated via Mendelian randomization (MR) where suitable exposure summary statistics were available, otherwise via direct variant–outcome associations. Complementary MR examined tissue-specific *cis*-eQTLs in brain and adipose tissues for genetically predicted ECS gene expression. Secondary outcomes included plasma/CSF biomarkers and UK Biobank neuroimaging phenotypes to explore mechanisms. Colocalization assessed shared causal variants.

**RESULTS:** After BH correction, *CNR2* rs4649124 was associated with increased VasD risk (P=0.049), increased all-cause dementia risk (P=0.051), and lower APP (P=0.088) and BDNF (P=0.083). *FAAH* rs324420 was associated with lower BDNF (P=0.037); *CNR1* rs12720071 with reduced bilateral hippocampal volume (P=0.095). Higher *CNR1* expression was associated with increased AD risk (cerebellum: P=0.001; cerebellar hemisphere: P=0.043) and higher APP (cerebellar hemisphere: P=0.062). Higher *DAGLB* expression was associated with increased all-cause dementia risk (visceral adipose: P=0.039; subcutaneous adipose: P=0.079; cerebellar hemisphere: P=0.079), higher BDNF (subcutaneous adipose: P=0.021), lower NfL (subcutaneous adipose: P=0.002; visceral adipose: P=0.011), lower bilateral amygdala volumes (subcutaneous adipose: P= 2.24 × 10□□, 0.026), and lower left ventral striatal volume (subcutaneous adipose: P=0.042). Higher cortical *MGLL* expression was associated with increased all-cause dementia risk (P=0.091). Colocalization did not support shared causal variants.

**DISCUSSION:** ECS variation showed dementia-related clinical, molecular, and neuroimaging associations, but absent colocalization limits causal inference. Findings warrant investigation.

## 1. Background

Dementia is a leading cause of disability and death worldwide, affecting over 55 million people globally, with Alzheimer’s disease (AD) accounting for the majority of cases, followed by vascular dementia (VasD) and other subtypes (1). Despite substantial research investment, disease-modifying treatments remain limited, highlighting the need to identify novel biological pathways involved in dementia pathogenesis (2,3).

Among the pathways under investigation, the endocannabinoid system (ECS) has emerged as a promising candidate (4,5). This endogenous lipid signalling system regulates synaptic transmission, neuroinflammation, neuronal survival, and neuroplasticity (6). Its core components include two G protein-coupled receptors — cannabinoid receptor type 1 (CB1, encoded by *CNR1*) and type 2 (CB2, encoded by *CNR2*) — and two primary ligands: anandamide (AEA) and 2-arachidonoylglycerol (2-AG). AEA is synthesised by N-acyl phosphatidylethanolamine phospholipase D (NAPEPLD) and degraded by fatty acid amide hydrolase (FAAH), while 2-AG is synthesised by diacylglycerol lipases alpha and beta (DAGLA, DAGLB) and degraded by monoacylglycerol lipase (MAGL) (7,8).

Growing evidence suggests ECS dysregulation may contribute to neurodegeneration. Post-mortem studies report altered CB1 and CB2 expression in AD brains, including CB2 upregulation in microglia surrounding amyloid plaques (9–11). Preclinical studies suggest ECS modulation can reduce amyloid-beta pathology, attenuate tau hyperphosphorylation, suppress neuroinflammation, and improve cognitive outcomes in neurodegeneration models. MAGL inhibition reduces amyloid-beta accumulation and neuroinflammation in AD mouse models, while FAAH inhibition exerts neuroprotective and anti-inflammatory effects and improves cognitive deficits in other models (12–14). Endocannabinoid signalling also intersects with neurotrophic pathways, including brain-derived neurotrophic factor (BDNF), and has been implicated in cerebrovascular regulation, suggesting relevance across dementia subtypes (15,16).

However, translating preclinical findings to humans remains challenging, as observational studies are susceptible to confounding and reverse causation. Mendelian randomization (MR) uses genetic variants as instrumental variables to estimate causal effects, offering an approach less susceptible to these biases (17,18). Although individual ECS components, including CB1, CB2, and FAAH, have been examined in AD and other neurodegenerative conditions (9–12,14), no study has systematically applied MR across the full ECS, spanning both receptors and metabolic enzymes, to investigate causal relevance for dementia risk and underlying biological mechanisms. This represents an important gap, given that ECS components operate as an integrated signalling network and that restricting analyses to single genes may fail to capture the breadth of ECS involvement in neurodegeneration.

Given evidence linking the ECS to neurodegeneration (9–12,14,15), we hypothesised that genetic variation influencing ECS function and expression may contribute to dementia risk and biological processes relevant to dementia, including amyloid and tau-related biology, neuroaxonal injury, glial activation and neurotrophic signalling, and structural brain alterations. This study aimed to characterise the contribution of ECS-related genetic variation to dementia using two complementary approaches. First, we systematically identified putatively functional variants within seven key ECS genes (*CNR1, CNR2, FAAH, DAGLA, DAGLB, NAPEPLD,* and *MGLL*), encoding the principal cannabinoid receptors and key synthetic and catabolic enzymes, and examined their relationship with AD, VasD, and all-cause dementia, using MR where suitable exposure summary statistics were available and direct variant–outcome associations otherwise. Second, we used MR to examine tissue-specific *cis*-expression quantitative trait loci (*cis*-eQTLs) for these genes in relation to the same dementia phenotypes, capturing genetically regulated gene expression rather than functional variant effects. Brain tissues were prioritised for their direct relevance to dementia pathophysiology; adipose tissues were additionally included given the ECS’s role in lipid metabolism and energy homeostasis (19), and growing evidence linking metabolic dysfunction to dementia risk (3,20). Secondary mechanistic outcomes included plasma protein levels, cerebrospinal fluid biomarkers, and UK Biobank neuroimaging phenotypes, allowing us to examine associations with molecular markers of amyloid- and tau-related biology, neuroaxonal injury, glial activation, neurotrophic signalling, and with structural brain phenotypes relevant to dementia. These analyses provided complementary mechanistic context for the primary dementia analyses by examining potential intermediate molecular and structural phenotypes linking ECS biology to dementia. Functional variant analyses of dementia outcomes constituted the main analysis of the study; tissue-specific *cis*-eQTL analyses were considered complementary. Results are presented and interpreted within this predefined analytical framework.

## 2. Methods

This study was conducted and reported in accordance with the STROBE-MR guidelines (21) for Mendelian randomization studies. An overview of the study design and analytical workflow is shown in Figure 1.

**Figure 1.**
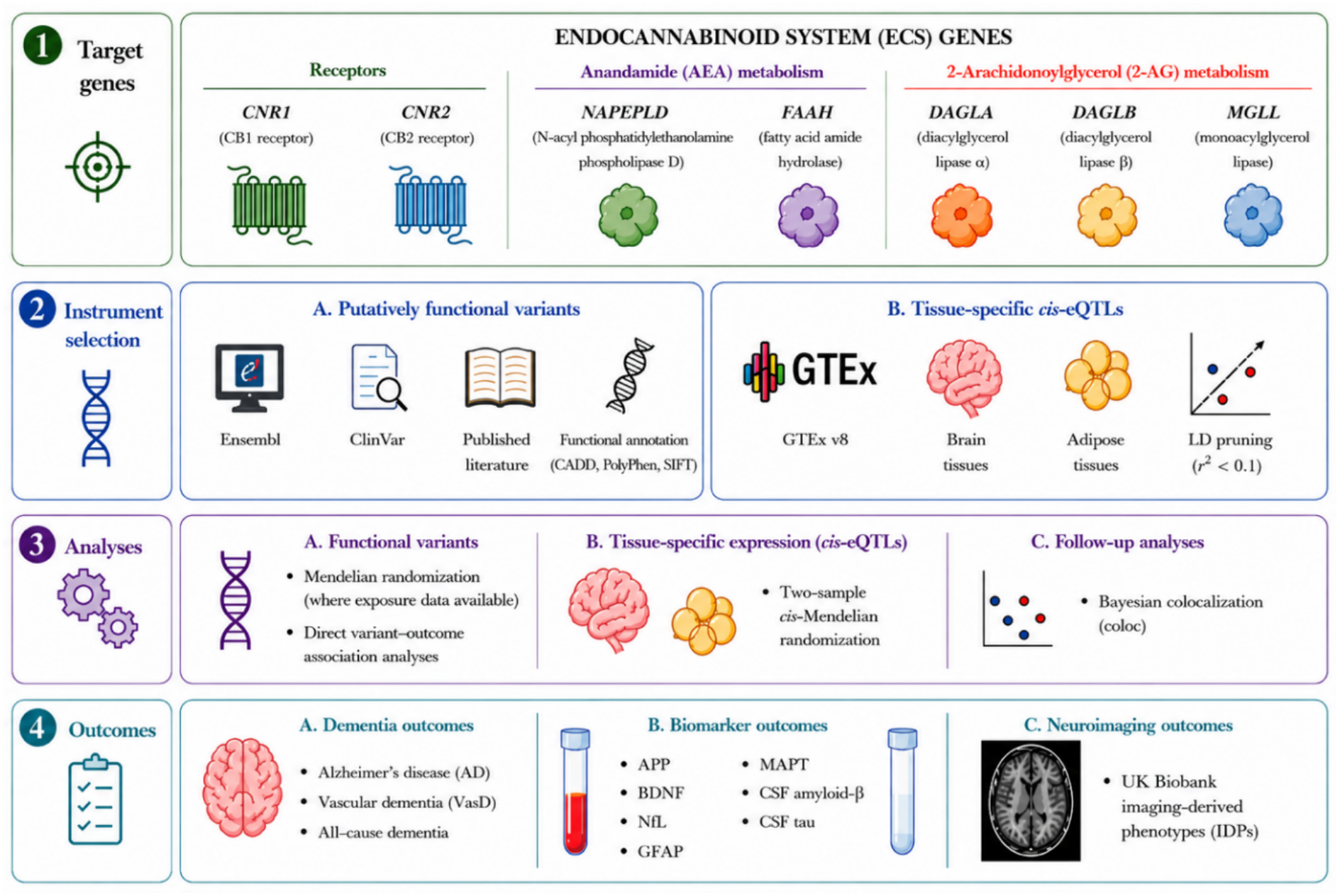
Study overview and analytical workflow. The endocannabinoid system (ECS) genes investigated in this study included receptor genes (*CNR1* and *CNR2*), genes involved in the anandamide (AEA) pathway (*NAPEPLD* and *FAAH*), and genes involved in the 2-arachidonoylglycerol (2-AG) pathway (*DAGLA*, *DAGLB*, and *MGLL*). Putatively functional variants were identified using Ensembl, ClinVar, published literature, and functional annotation resources. Tissue-specific *cis*-expression quantitative trait loci (*cis*-eQTLs) were obtained from GTEx v8 and analysed in brain and adipose tissues following linkage disequilibrium pruning (r² < 0.1). Functional variants were investigated using Mendelian randomization (MR) where suitable exposure summary statistics were available, or through direct variant–outcome association analyses where these were unavailable. Tissue-specific expression analyses were conducted using two-sample *cis*-MR, followed by Bayesian colocalization analyses. Outcomes included Alzheimer’s disease, vascular dementia, all-cause dementia, plasma and cerebrospinal fluid biomarkers, and UK Biobank neuroimaging-derived phenotypes (IDPs).

### 2.1 Instrument selection

#### 2.1.1 Selection of putatively functional variants

Putatively functional variants were prioritised to provide biologically interpretable genetic instruments (18). Variants in ECS genes were identified using a combination of bioinformatic resources and literature review. Supplementary Table S1 provides the detailed functional annotation of all selected variants. Coding variants, including missense, stop-gained, frameshift, in-frame insertions/deletions, and splice-site variants, were first extracted from Ensembl. For missense variants, available bioinformatic annotations, including Combined Annotation Dependent Depletion (CADD), PolyPhen, and SIFT scores, were used to assess potential functional impact. Additional information on evolutionary conservation and clinical annotation was obtained from GERP scores and ClinVar classifications. To identify additional variants with potential biological relevance, ClinVar and published literature were also reviewed. Consequently, the final variant set included missense, synonymous, and regulatory variants with evidence of functional, regulatory, or phenotypic relevance. Where available, variants with a minor allele frequency (MAF) > 0.01 were retained. Pairwise linkage disequilibrium (LD) between functional variants within genes containing more than one eligible variant was assessed using LDlink based on the European reference population from the 1000 Genomes Project (Supplementary Table S2). Variants in low LD were considered independent and analysed separately. Where functional variants were in complete LD (r² = 1), they were considered to represent the same underlying genetic signal, and a single representative variant was retained for downstream analyses. All selected variants were subsequently checked for availability across the outcome datasets used in this study (22–26).

#### 2.1.2 Selection of cis-eQTLs

*Cis*-eQTLs for ECS genes were obtained from the Genotype-Tissue Expression (GTEx) project (version 8) (27). The list of genes and availability of *cis*-eQTLs in brain and adipose tissues is provided in Supplementary Table S19. Brain and adipose tissues were included based on their biological relevance (19) and the availability of *cis*-eQTL signals for ECS genes. Further details of GTEx *cis*-eQTL generation, variant filtering and instrument selection are provided in Supplementary Methods M1. Supplementary Table S19 summarises *cis*-eQTL availability, while Supplementary Table S19a lists the LD-independent *cis*-eQTL genetic instruments retained after LD clumping (r² < 0.1) and used in the MR analyses.

### 2.2 Outcome datasets

#### 2.2.1 Primary dementia outcomes

Publicly available summary statistics from genome-wide association studies (GWASs) conducted in European ancestry populations were used, including Alzheimer’s disease (n = 94,437; cases = 35,274, clinically diagnosed or autopsy-confirmed) (22), vascular dementia (n = 466,606; cases = 3,892, ascertained using validated clinical diagnostic criteria, with ICD-10 codes used in UK Biobank) (23), and all-cause dementia (n = 466,606; cases = 44,009, ascertained using validated clinical diagnostic criteria, with proxy dementia based on parental history used in UK Biobank) (Supplementary Tables S3) (23).

#### 2.2.2 Secondary mechanistic outcomes

Genetic associations for biomarkers and neuroimaging endophenotypes were obtained from previously published GWAS studies to investigate potential biological mechanisms underlying dementia associations. These included cerebrospinal fluid (CSF) tau and amyloid-beta levels (n = 13,116) (24); Olink-derived measures of circulating protein abundance in UK Biobank participants (n ≤ 54,219) (25), specifically neurofilament light chain (NfL), glial fibrillary acidic protein (GFAP), amyloid precursor protein (APP), brain-derived neurotrophic factor (BDNF), and microtubule-associated protein tau (MAPT), selected a priori for their relevance to dementia-related pathways, including neuroaxonal injury, glial activation, amyloid- and tau-related biology, and neurotrophic signalling; and neuroimaging-derived phenotypes (IDPs) from the UK Biobank (n ≈ 33,000) (26). Additional GWAS datasets were included for colocalization analyses where relevant, including oleoylethanolamide levels (n = 8,243) (28) (Supplementary Tables S3 and S4).

### 2.3 Genetic analyses

#### 2.3.1 Mendelian randomization analyses

Two-sample MR was used to estimate the potential causal effects of genetic variation in seven individual ECS components — the CB1 and CB2 receptors (encoded by *CNR1* and *CNR2*) and five metabolic enzymes (encoded by *FAAH*, *DAGLA*, *DAGLB*, *NAPEPLD*, and *MGLL*) — on dementia outcomes and secondary mechanistic outcomes (Supplementary Tables S3 and S4). *cis*-MR was applied by restricting instruments to variants located within or near the gene of interest to reduce horizontal pleiotropy. Tissue-specific *cis*-eQTL instruments were derived from GTEx. These were used to test associations between genetically predicted ECS gene expression and the primary dementia outcomes. Tissue-specific analyses followed a hierarchical framework, whereby secondary mechanistic analyses (plasma/CSF biomarkers and UK Biobank neuroimaging-derived phenotypes) were performed only for tissue-specific *cis*-eQTLs that remained significantly associated with at least one primary dementia outcome after correction for multiple testing. Results were evaluated at both 5% and 10% FDR thresholds, with the 5% threshold representing the more stringent level of evidence. All associations meeting an FDR ≤10% were taken forward for further mechanistic analyses. The inverse-variance weighted (IVW) method was used as the primary analysis. Where only one genetic instrument was available, the Wald ratio was used. MR-Egger and Mendelian Randomization Pleiotropy RESidual Sum and Outlier (MR-PRESSO) were not performed because the number of independent instrumental variants was limited in most analyses, reducing the reliability of pleiotropy assessment and outlier detection (18,29). Analyses were conducted using the TwoSampleMR package (version 0.6.3). Variant harmonisation was performed to ensure alignment of effect alleles across exposure and outcome datasets.

#### 2.3.2 Variant–outcome association analyses

For functional variants, MR analyses were performed when suitable summary statistics for a relevant exposure associated with the genetic variant were available. When such exposure summary statistics were unavailable, direct variant–outcome association analyses were performed instead. The functional variants, their associated exposure traits where available, and supporting evidence are detailed in Supplementary Table S1. Where variants were unavailable in specific outcome datasets and no suitable proxy variant could be identified, analyses were restricted to the remaining eligible variant(s). For genes containing multiple independent functional variants, gene-level evidence was summarised by combining variant-specific *P*-values using Fisher’s method (30). For *CNR2*, rs4649124 was retained as the representative functional variant because it was in complete linkage disequilibrium (r² = 1) with rs35761398 (Supplementary Table S2). Consequently, gene-level *P*-value combination was not performed for *CNR2*. A second *CNR2* functional variant (rs61996280) was excluded because neither the variant nor a suitable proxy was available in the outcome datasets.

Dementia outcomes, neurobiological biomarkers, and neuroimaging-derived phenotypes were examined using the most appropriate analytical approach according to data availability (Supplementary Tables S3–S4). When variants were not present in outcome datasets, LD proxies were identified using LDlink based on the European reference population from the 1000 Genomes Project, with proxies selected where suitable (r² ≥ 0.8). Additional gene-specific analytical details are provided in the Supplementary Materials (M2).

### 2.4 Colocalization analyses

Bayesian colocalization analysis (coloc, version 5.2.3) was performed to assess whether associations were driven by shared causal variants (31,32). Prior probabilities of p1 = 1 × 10□□, p2 = 1 × 10□□, and p12 = 1 × 10□□ were used, with the lower p12 value providing a more conservative prior for a shared causal variant. Posterior probabilities were estimated for five hypotheses (H0–H4), including a shared causal variant (PP.H4) and distinct causal variants (PP.H3). Evidence of colocalization was defined as PP.H4 + PP.H3 > 0.5 and PP.H4 / (PP.H4 + PP.H3) > 0.5, indicating sufficient evidence for association with both traits and greater support for a shared rather than distinct causal variant (31). Values not meeting these criteria were interpreted as lack of support for a shared causal variant. For selected findings, including *FAAH* and *DAGLB*, colocalization analyses were performed using *cis*-eQTL datasets and, where appropriate, GWAS-derived proxy traits. For *FAAH*, two colocalization analyses were performed: one using *FAAH cis*-eQTLs across multiple brain tissues and adipose subcutaneous tissue, and another using a GWAS of oleoylethanolamide (OEA), which was selected as a proxy of *FAAH* activity because rs324420 is strongly associated with circulating OEA levels and suitable GWAS summary statistics for anandamide (AEA) were not available (28). For *DAGLB*, colocalization analyses were performed using *DAGLB cis*-eQTLs. Further gene-specific methodological details are provided in the Supplementary Materials (M3).

### 2.5 Multiple testing correction

Multiple testing correction was applied separately for each ECS gene because the study evaluated each ECS component individually as a candidate target using distinct genetic instruments. Although these genes participate in the same endocannabinoid pathway, they have different biological roles and molecular functions. Within each gene, the primary dementia outcomes and secondary mechanistic outcomes were treated as separate families of hypotheses and corrected independently using the Benjamini–Hochberg (BH) false discovery rate (FDR) procedure. BH-adjusted P-values were calculated using the p.adjust(method = "fdr") function in R. Findings meeting the 5% and 10% FDR thresholds are reported where relevant (33).

## 3. Results

The principal findings are summarised in Table 1, with detailed results presented in the Supplementary Tables and Figures. Functional variant analyses constituted the primary analyses of the study and are presented first. Within each analysis, genes are ordered according to the extent of significant findings to facilitate interpretation. These are followed by the complementary tissue-specific *cis*-eQTL analyses. Colocalization analyses were subsequently performed to investigate whether selected associations reflected shared causal variants.

**Table 1.**
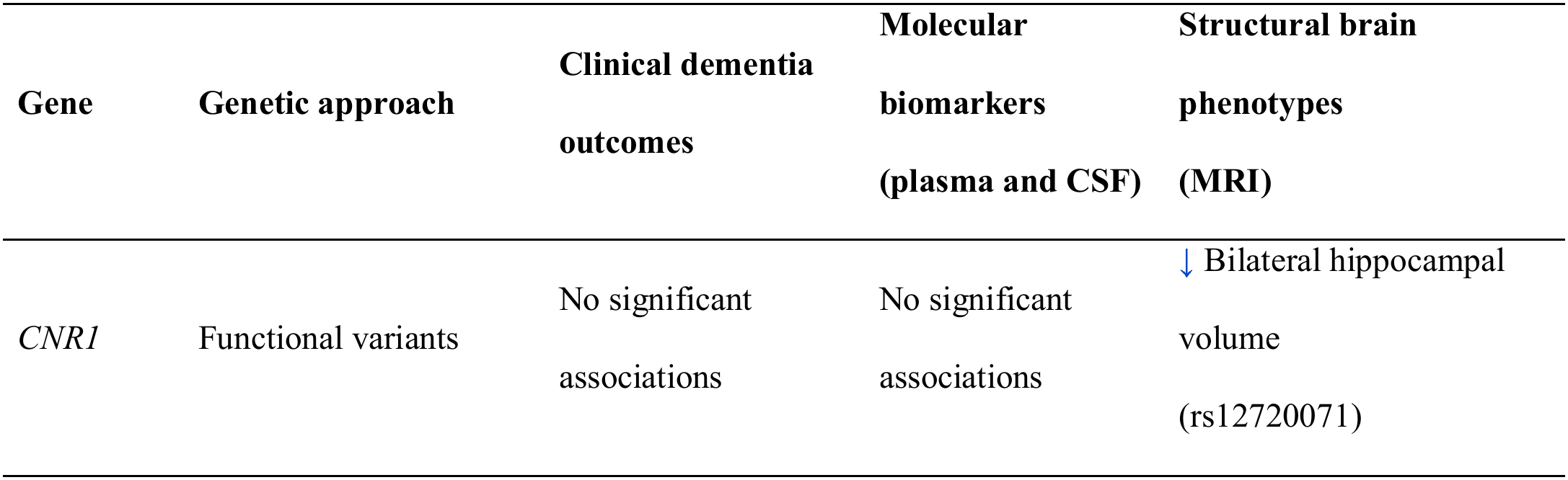
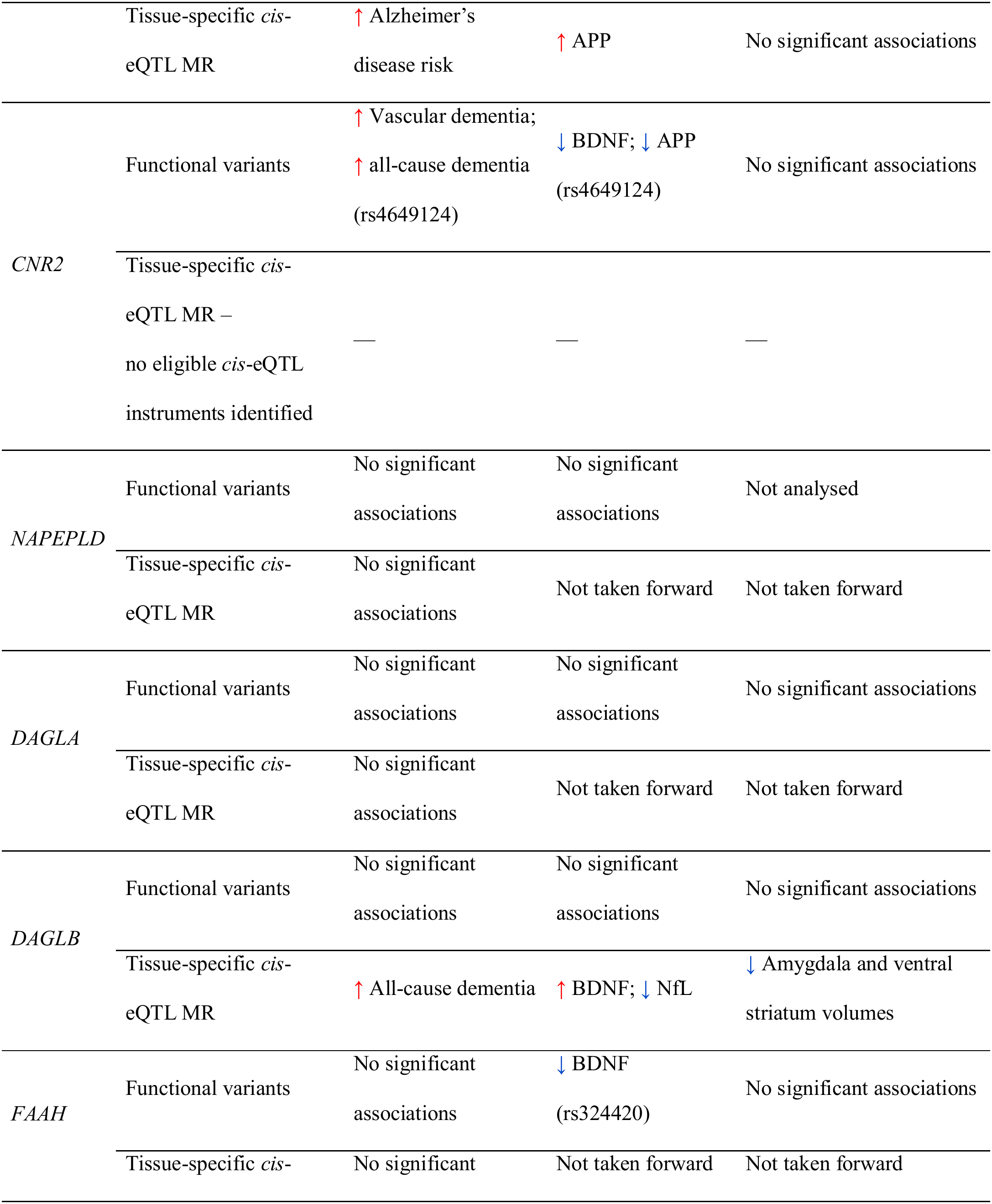
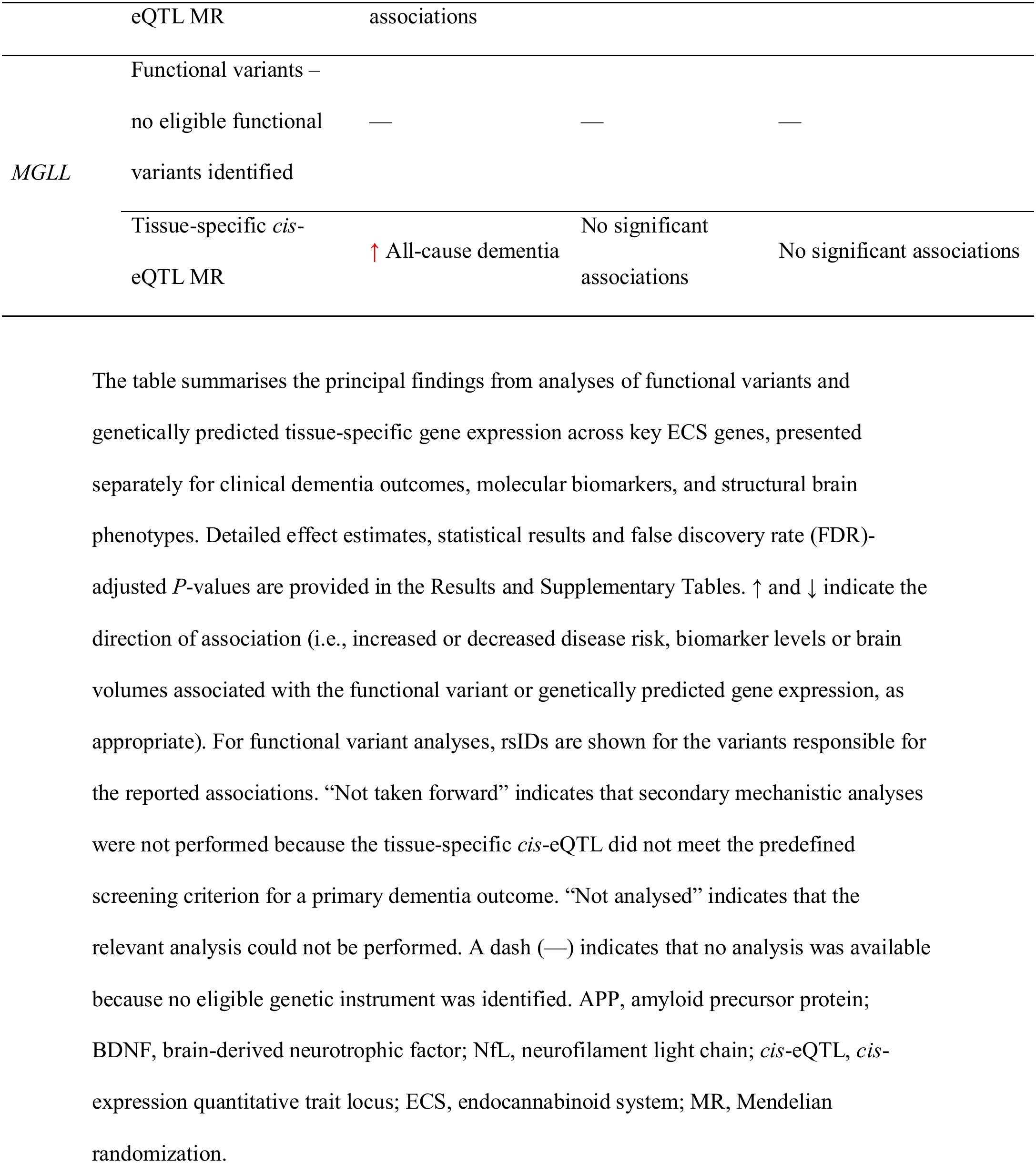
Summary of key findings across functional variant and tissue-specific expression analyses of ECS genes.

| Gene | Genetic approach | Clinical dementia outcomes | Molecular biomarkers (plasma and CSF) | Structural brain phenotypes (MRI) |
|---|---|---|---|---|
| *CNR1* | Functional variants | No significant associations | No significant associations | ↓ Bilateral hippocampal volume (rs12720071) |
|  | Tissue-specific *cis*-eQTL MR | ↑ Alzheimer’s disease risk | ↑ APP | No significant associations |
| *CNR2* | Functional variants | ↑ Vascular dementia; ↑ all-cause dementia (rs4649124) | ↓ BDNF; ↓ APP (rs4649124) | No significant associations |
|  | Tissue-specific *cis*-eQTL MR – no eligible *cis*-eQTL instruments identified | — | — | — |
| *NAPEPLD* | Functional variants | No significant associations | No significant associations | Not analysed |
|  | Tissue-specific *cis*-eQTL MR | No significant associations | Not taken forward | Not taken forward |
| *DAGLA* | Functional variants | No significant associations | No significant associations | No significant associations |
|  | Tissue-specific *cis*-eQTL MR | No significant associations | Not taken forward | Not taken forward |
| *DAGLB* | Functional variants | No significant associations | No significant associations | No significant associations |
|  | Tissue-specific *cis*-eQTL MR | ↑ All-cause dementia | ↑ BDNF; ↓ NfL | ↓ Amygdala and ventral striatum volumes |
| *FAAH* | Functional variants | No significant associations | ↓ BDNF (rs324420) | No significant associations |
|  | Tissue-specific *cis*-eQTL MR | No significant associations | Not taken forward | Not taken forward |
| *MGLL* | Functional variants – no eligible functional variants identified | — | — | — |
|  | Tissue-specific *cis*-eQTL MR | ↑ All-cause dementia | No significant associations | No significant associations |

The table summarises the principal findings from analyses of functional variants and genetically predicted tissue-specific gene expression across key ECS genes, presented separately for clinical dementia outcomes, molecular biomarkers, and structural brain phenotypes. Detailed effect estimates, statistical results and false discovery rate (FDR)-adjusted *P*-values are provided in the Results and Supplementary Tables. ↑ and ↓ indicate the direction of association (i.e., increased or decreased disease risk, biomarker levels or brain volumes associated with the functional variant or genetically predicted gene expression, as appropriate). For functional variant analyses, rsIDs are shown for the variants responsible for the reported associations. “Not taken forward” indicates that secondary mechanistic analyses were not performed because the tissue-specific *cis*-eQTL did not meet the predefined screening criterion for a primary dementia outcome. “Not analysed” indicates that the relevant analysis could not be performed. A dash (—) indicates that no analysis was available because no eligible genetic instrument was identified. APP, amyloid precursor protein; BDNF, brain-derived neurotrophic factor; NfL, neurofilament light chain; *cis*-eQTL, *cis*-expression quantitative trait locus; ECS, endocannabinoid system; MR, Mendelian randomization.

### 3.1 Functional variant analyses

#### 3.1.1 CNR2

The representative *CNR2* missense variant rs4649124 was associated with an increased risk of vascular dementia (β = 0.07, 95% CI = [0.01, 0.12], *P* = 0.016; BH-adjusted *P* = 0.049) and all-cause dementia (β = 0.03, 95% CI = [2 × 10□³, 0.05], *P* = 0.034; BH-adjusted *P* = 0.051). The association with vascular dementia remained significant after correction at the 5% FDR threshold, whereas the association with all-cause dementia remained below the 10% FDR threshold. No evidence of an association with Alzheimer’s disease was observed (Supplementary Table S5).

Among the secondary mechanistic outcomes, rs4649124 was associated with reduced circulating BDNF levels (β = −0.02, 95% CI = [−0.03, −4 × 10□ ³], *P* = 0.012; BH-adjusted *P* = 0.083) and APP levels (β = −0.02, 95% CI = [−0.03, −2 × 10□³], *P* = 0.025; BH-adjusted *P* = 0.088). Both associations remained below the 10% FDR threshold but not the 5% threshold, whereas no evidence of associations was observed for Aβ, tau, GFAP, MAPT, or NfL (Supplementary Table S5).

No associations were observed with neuroimaging-derived phenotypes (Supplementary Table S6).

#### 3.1.2 FAAH

The *FAAH* missense variant rs324420 was associated with reduced circulating BDNF levels (Wald ratio, β = −0.05, 95% CI = [−0.09, −0.02], *P* = 0.005; BH-adjusted *P* = 0.037), remaining significant after correction at the 5% FDR. No evidence of association was observed for APP, Aβ, tau, GFAP, MAPT, or NfL (Supplementary Table S7). No neuroimaging-derived phenotypes remained associated with rs324420 after correction (Supplementary Table S8). The variant also showed no evidence of association with Alzheimer’s disease, vascular dementia, or all-cause dementia (Supplementary Table S7).

#### 3.1.3 CNR1

For neuroimaging-derived phenotypes, gene-level analyses identified associations between *CNR1* and reduced right and left hippocampal volumes, which remained below the 10% FDR threshold, although not the 5% threshold (Fisher combined *P* = 0.007 and 0.011, respectively; BH-adjusted *P* = 0.095 for both; Supplementary Table S11). These associations were driven by the missense variant rs12720071, which was associated with reduced right hippocampal volume (β = −0.04, 95% CI = [−0.07, −0.02], *P* = 0.002; BH-adjusted *P* = 0.076) and reduced left hippocampal volume (β = −0.04, 95% CI = [−0.07, −0.01], *P* = 0.004; BH-adjusted *P* = 0.076) (Supplementary Table S11a). Gene-level analyses showed no evidence of association with Alzheimer’s disease, vascular dementia, all-cause dementia, or plasma and CSF biomarkers (Supplementary Table S10). Individual variant associations with the primary dementia and secondary mechanistic outcomes are presented in Supplementary Table S10a.

#### 3.1.4 Other ECS functional variants (DAGLB, DAGLA, NAPEPLD and MGLL)

No robust evidence of association was identified for the remaining ECS functional variants across the primary dementia outcomes, secondary mechanistic biomarkers, or neuroimaging-derived phenotypes. Specifically, the *DAGLB* missense variant rs1055430 showed no evidence of association across all analysed outcomes (Supplementary Tables S12 and S13). Likewise, neither Mendelian randomization nor functional variant analyses identified robust associations for *DAGLA* (Supplementary Tables S14–S17; Supplementary Figures S1 and S2). The *NAPEPLD* missense-regulatory variant rs3181009 showed no evidence of association with the analysed dementia or biomarker outcomes. Neuroimaging analyses were not performed because neither the variant nor a suitable proxy was available in the UK Biobank IDPs GWAS (Supplementary Table S18). No functional variants meeting the predefined selection criteria were identified for *MGLL*; therefore, *MGLL* was evaluated only in the complementary *cis*-eQTL analyses.

The overall pattern of associations across the evaluated functional variants is summarised in Figure 2, illustrating the direction and magnitude of effect estimates for the primary dementia outcomes and the secondary plasma and CSF biomarker outcomes.

**Figure 2.**
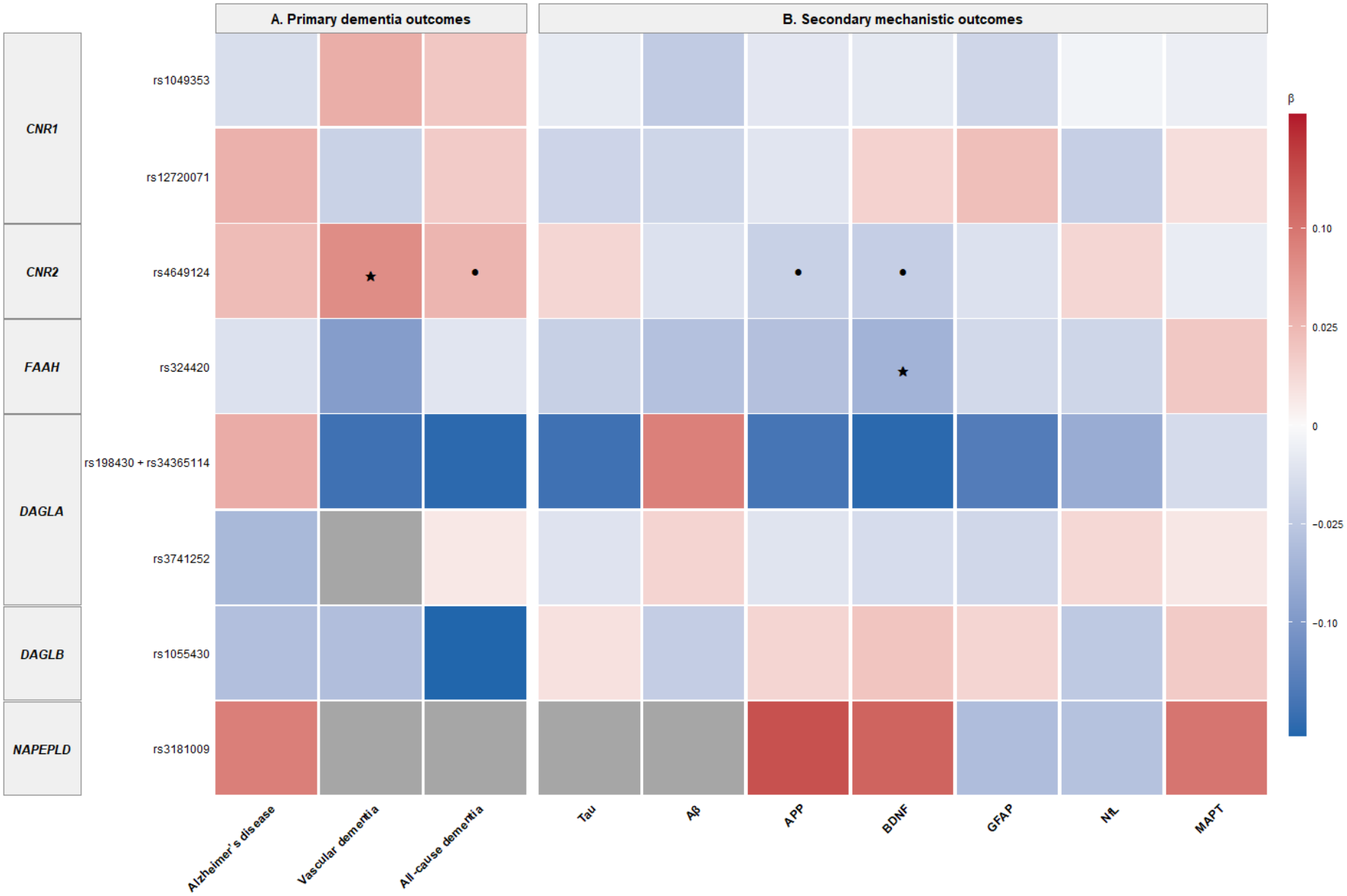
Overview of functional variant associations across primary dementia outcomes and secondary plasma and CSF biomarker outcomes. Heatmap illustrating the direction and magnitude of Mendelian randomization effect estimates (β) for the evaluated functional variants across the primary dementia outcomes (Alzheimer’s disease, vascular dementia and all-cause dementia) and secondary plasma and CSF biomarker outcomes (tau, Aβ, APP, BDNF, GFAP, NfL and MAPT). Red indicates positive associations and blue indicates negative associations. Black stars indicate associations remaining significant after Benjamini–Hochberg correction at a 5% false discovery rate (FDR), and black circles indicate associations meeting the 10% FDR threshold but not the 5% threshold. Grey cells indicate outcomes that were not analysed. *MGLL* is not shown because no eligible functional variant satisfied the predefined inclusion criteria.

### 3.2 Tissue-specific expression analyses

Tissue-specific *cis*-eQTL analyses identified evidence of association for *CNR1* and *DAGLB* with the primary dementia outcomes. More limited evidence was observed for *MGLL*, whereas *FAAH*, *DAGLA*, and *NAPEPLD* showed no robust associations.

#### 3.2.1 CNR1

Genetically predicted higher *CNR1* expression in the cerebellum and cerebellar hemisphere was associated with an increased risk of Alzheimer’s disease (IVW, β = 0.18, 95% CI = [0.08, 0.28], *P* = 3 × 10□□; BH-adjusted *P* = 0.001; and β = 0.16, 95% CI = [0.02, 0.30], *P* = 0.020; BH-adjusted *P* = 0.043, respectively) (Supplementary Table S20). Higher genetically predicted *CNR1* expression in the cerebellar hemisphere was also associated with higher APP levels at the 10% FDR threshold (IVW, β = 0.06, 95% CI = [3 × 10□³, 0.12], *P* = 0.038; BH-adjusted *P* = 0.062). No evidence of association was observed with vascular dementia, all-cause dementia, other plasma or CSF biomarkers, or neuroimaging-derived phenotypes (Supplementary Tables S20–S22; Supplementary Figures S3–S5).

#### 3.2.2 DAGLB

Genetically predicted higher *DAGLB* expression showed evidence of association with all-cause dementia, circulating biomarkers, and neuroimaging-derived phenotypes. Higher expression in visceral adipose tissue was associated with an increased risk of all-cause dementia (IVW, β = 0.05, 95% CI = [0.02, 0.08], *P* = 0.004; BH-adjusted *P* = 0.039). At the 10% FDR threshold, similar associations were observed for subcutaneous adipose tissue (IVW, β = 0.02, 95% CI = [3 × 10□ ³, 0.05], *P* = 0.023; BH-adjusted *P* = 0.079) and the cerebellar hemisphere (IVW, β = 0.04, 95% CI = [0.01, 0.08], *P* = 0.024; BH-adjusted *P* = 0.079) (Supplementary Table S23; Supplementary Figure S6).

Among the biomarker outcomes, genetically predicted higher *DAGLB* expression in subcutaneous adipose tissue was associated with higher BDNF levels (IVW, β = 0.02, 95% CI = [4 × 10□ ³, 0.03], *P* = 0.007; BH-adjusted *P* = 0.021). Higher expression in both subcutaneous and visceral adipose tissues was associated with lower NfL levels (IVW, β = −0.02, 95% CI = [−0.03, −0.01], *P* = 6.94 × 10□□; BH-adjusted *P* = 0.002; and β = −0.02, 95% CI = [−0.04, −0.01], *P* = 0.008; BH-adjusted *P* = 0.011, respectively). At the 10% FDR threshold, lower NfL levels were also observed for cerebellar hemisphere expression (IVW, β = −0.02, 95% CI = [−0.05, 1 × 10□³], *P* = 0.011; BH-adjusted *P* = 0.060). No evidence of association was observed for Aβ, tau, APP, GFAP, or MAPT (Supplementary Table S24; Supplementary Figure S7).

Neuroimaging analyses showed multiple associations between genetically predicted higher *DAGLB* expression and lower amygdala and ventral striatal volumes across several tissues. Higher expression in subcutaneous adipose tissue was associated with lower left amygdala volume (IVW, β = −0.04, 95% CI = [−0.05, −0.03], *P* = 1.32 × 10^-8^; BH-adjusted *P* = 2.24 ×

10□□), lower right amygdala volume (IVW, β = −0.02, 95% CI = [−0.03, −0.01], *P* = 0.003; BH-adjusted *P* = 0.026), and lower left ventral striatum grey matter volume (IVW, β = −0.02, 95% CI = [−0.03, −5 × 10□□³], *P* = 0.007; BH-adjusted *P* = 0.042). Higher expression in visceral adipose tissue was also associated with lower left amygdala volume (IVW, β =

−0.03, 95% CI = [−0.05, −0.02], *P* = 9.41 × 10^-5^; BH-adjusted *P* = 0.002) and lower left ventral striatum grey matter volume (IVW, β = −0.03, 95% CI = [−0.04, −0.01], *P* = 0.002; BH-adjusted *P* = 0.013). In addition, higher expression in the cerebellar hemisphere was associated with lower left amygdala volume (IVW, β = −0.04, 95% CI = [−0.06, −0.02], *P* = 9.82 × 10^-4^; BH-adjusted *P* = 0.017; Supplementary Table S25; Supplementary Figure S8).

#### 3.2.3 Other ECS genes (FAAH, DAGLA, NAPEPLD and MGLL)

No evidence of association was observed between genetically predicted *FAAH*, *DAGLA*, or *NAPEPLD* expression and Alzheimer’s disease, vascular dementia, or all-cause dementia after correction across the evaluated tissues (Supplementary Tables S26–S28; Supplementary Figures S9–S11). In contrast, at the 10% FDR threshold, genetically predicted higher *MGLL* expression in the cortex was associated with an increased risk of all-cause dementia (Wald ratio, β = 0.19, 95% CI = [0.04, 0.34], *P* = 0.011; BH-adjusted *P* = 0.091). However, no evidence of association was observed with the evaluated plasma and CSF biomarkers or neuroimaging-derived phenotypes (Supplementary Tables S29–S31; Supplementary Figure S12).

### 3.3 Colocalization analyses

Across both the functional variant and tissue-specific expression analyses, colocalization analyses provided no evidence to support shared causal variants underlying the observed associations.

For the functional variant analyses, no evidence of colocalization was observed between the OEA GWAS signal (used as a proxy for *FAAH* activity) and the BDNF signal. Similarly, no evidence of colocalization was observed between *FAAH cis*-eQTLs and the BDNF signal (Supplementary Table S9 and Supplementary Figure S13P–Z). Although PP.H4 exceeded PP.H3 in both analyses, the OEA-based analysis showed PP.H4 = 0.217 and PP.H3 = 0.065, while the *FAAH cis*-eQTL analyses showed PP.H4 values ranging from 0.009 to 0.015 and PP.H3 values ranging from 0.001 to 0.005. These findings did not meet the predefined criteria for colocalization.

Similarly, tissue-specific colocalization analyses showed no evidence of shared causal variants between the *cis*-eQTL and GWAS signals for any of the evaluated associations. Posterior probabilities supporting colocalization were consistently low (PP.H4 < 0.05 across all analyses) (Supplementary Table S32 and Supplementary Figure S13A–Y).

## 4. Discussion

This study provides the first systematic MR investigation of the ECS in relation to dementia risk, spanning receptor genes (*CNR1*, *CNR2*) and metabolic enzymes (*FAAH*, *DAGLA*, *DAGLB*, *NAPEPLD*, *MGLL*). By integrating functionally predicted coding variants, particularly missense variants, with complementary *cis*-eQTLs, we examined functional variation and genetically regulated ECS expression in relation to clinical dementia outcomes and selected biological processes, including amyloid- and tau-related biology, neuroaxonal injury, glial activation and neurotrophic signalling, and brain structure. Three principal findings emerged: functional variant analyses identified evidence that selected ECS coding variants may influence dementia risk and neurobiological traits; *cis*-eQTL analyses suggested genetically predicted ECS expression may contribute to selected dementia-related pathways; and although several biologically plausible associations survived correction, none was supported by colocalization, limiting causal inference. Overall, specific ECS components may contribute to selected pathways relevant to dementia, with limited evidence for a broad causal role.

### 4.1 CNR2 variants and dementia risk

The most consistent finding centres on *CNR2*. The missense variant rs4649124 was associated with increased vascular dementia risk (β = 0.07, 95% CI = [0.01, 0.12]; BH-adjusted *P* = 0.049), surviving the 5% FDR, with associations for all-cause dementia and reduced APP and BDNF only at 10% FDR. CB2 receptors are expressed predominantly on microglia and peripheral immune cells, where activation suppresses pro-inflammatory cytokine release and modulates microglial activation, establishing CB2 as a key regulator (34–36). CB2 signalling has been implicated in cerebrovascular inflammation and neurovascular function (37). CB2-deficient mouse models show loss of CB2 alters amyloid burden and dysregulates microglial responses in APP transgenic models, though amyloid effect direction has varied across models, likely reflecting transgenic background, disease stage, and microglial function examined (38–40).

The finding that CB2 loss disrupts microglia–amyloid relationships across models supports a role for CB2 in APP metabolism. The lower-APP finding is consistent, though modest correction strength warrants caution. No association was observed with AD specifically, despite AD being the largest all-cause dementia component— possibly reflecting the larger sample size of the all-cause dementia GWAS (23), and because APP levels reflect broader amyloid metabolism not specific to AD (22). The concurrent vascular dementia association raises the possibility that the all-cause signal partly reflects vascular/mixed-pathology rather than AD pathology, consistent with CB2’s cerebrovascular role (37,41). Together, these findings suggest *CNR2* variation may influence dementia risk through vascular and amyloid-related pathways, though relative contribution remains unclear. Caution is warranted given rs4649124’s modest functional annotation, reliance on a single variant, and absent colocalization support; associations are regarded as hypothesis-generating.

### 4.2 FAAH variation and circulating BDNF

The *FAAH* missense variant rs324420 (P129T; Supplementary Table S1), which reduces FAAH stability and is predicted to increase AEA (42), was associated with lower circulating BDNF after correction — counterintuitive given AEA’s generally neuroprotective role (43,44). CB1 activation downstream of AEA has been linked to BDNF regulation via PI3K/Akt/mTORC1 and CREB-mediated transcription (45); chronic AEA elevation may cause CB1 desensitisation, with complex downstream neurotrophic effects. Given BDNF’s role in neuronal survival and its reduction in AD (46,47), this supports a potential neurotrophic mechanism, though caution is warranted given the lack of colocalization support. No association was found between rs324420 and dementia outcomes, possibly reflecting single-SNP power, though any true effect may be modest or indirect. Nominal associations with white matter hyperintensity and amygdala volume did not survive correction. Colocalization showed weak preferential support without reaching thresholds, possibly reflecting limited power rather than absent biological relationship.

### 4.3 CNR1 and hippocampal structure

For *CNR1*, functional variant analyses found no dementia or biomarker associations. The association between rs12720071 and reduced bilateral hippocampal volume — a region of established high CB1 density (48,49) — is suggestive only, significant at the 10% rather than 5% FDR, but provides structural support for *CNR1*’s neurobiological relevance.

Complementary *cis*-eQTL analyses linked higher *CNR1* expression in cerebellar regions to increased AD risk and higher APP. While the cerebellum has historically been considered relatively spared in AD (50,51), emerging evidence suggests a role in disease-related molecular changes (52,53) and CB1 receptors are highly expressed in cerebellar neurons (54), providing relevant context. However, *cis*-eQTL analyses assess genetically predicted expression differences within tissues rather than absolute regional differences, so the observed associations cannot be directly attributed to high cerebellar CB1 expression. Experimental studies suggest CB1 signalling may influence APP processing (55). Lack of colocalization support limits causal inference, and findings should be interpreted cautiously.

### 4.4 DAGLB: functional variant and tissue-specific expression findings

The *DAGLB* functional variant rs1055430 showed no association with any outcome. In contrast, *c*is-eQTL analyses provided the most extensive evidence across genes: higher genetically predicted adipose *DAGLB* expression was associated with increased all-cause dementia risk, higher BDNF, lower NfL, and lower amygdala and ventral striatal volumes; higher cerebellar hemisphere expression was associated with increased all-cause dementia risk and lower left amygdala volume. *DAGLB* is the primary 2-AG synthesising enzyme in microglia (36,56), and microglial endocannabinoid signalling has been implicated in neuroinflammatory processes relevant to neurodegeneration (36,57), offering a plausible biological basis. Direction is not straightforward: higher *DAGLB* expression was linked to both increased dementia risk and lower NfL (generally a marker of reduced neurodegeneration), suggesting complex, tissue-specific rather than uniformly protective/harmful pathways. Colocalization did not support shared causal variants, generally supporting genetic association with *DAGLB* expression but not with outcome; findings should be considered hypothesis-generating.

### 4.5 MGLL cortical expression and dementia risk

Complementary eQTL analyses linked higher cortical *MGLL* expression to increased all-cause dementia risk (10% FDR). *MGLL* encodes MAGL, key to 2-AG degradation and ECS tone/inflammatory regulation (58). Preclinical studies show MAGL inhibition reduces neuroinflammation and amyloid pathology (58,59), and present findings are directionally consistent. However, no biomarker or neuroimaging associations were observed, and colocalization did not support a shared causal variant, so findings should be considered hypothesis-generating.

### 4.6 Convergent ECS–BDNF and APP signals across genes

A recurring signal concerns circulating BDNF, associated with three ECS genes across both analytical strata: *CNR2* rs4649124 (10% FDR), *FAAH* rs324420 (5% FDR), and *DAGLB* expression in subcutaneous adipose tissue (5% FDR). This convergence strengthens the possibility that ECS signalling is genuinely linked to neurotrophic pathways in dementia, consistent with prior cannabinoid–BDNF evidence (15,45).

Direction was not uniform: *CNR2* and *FAAH* variants were associated with lower BDNF, while higher *DAGLB* adipose expression was associated with higher BDNF. This may reflect differing mechanisms — *CNR2/FAAH* arise from coding variants affecting receptor/enzyme function directly, whereas *DAGLB* reflects predicted expression in peripheral tissue, potentially a distinct adipose-mediated route rather than central neurotrophic signalling.

Alternatively, the ECS–BDNF relationship may not be linear or uniformly directional across system nodes, consistent with the broader complexity noted above. Given the absence of colocalization support, this should be regarded as hypothesis-generating and warrants further investigation using multi-tissue expression and functional data.

A related pattern was observed for APP: CNR2 rs4649124 was associated with lower APP (10% FDR), while higher CNR1 cerebellar hemisphere expression was associated with higher APP (10% FDR). Neither reached 5% FDR, and opposing directions again distinguish a receptor-level variant from a tissue-specific expression signal. Both are consistent with the biological rationale discussed above, but given weaker statistical support relative to BDNF, this is a tentative secondary observation rather than firm evidence of a shared APP pathway.

### 4.7 Null findings and interpretation

*DAGLA* and *NAPEPLD* showed no robust associations after correction, likely reflecting limited instrument availability, modest effect sizes, and ECS signalling complexity across tissues. The absence of association between *FAAH* expression and dementia outcomes, despite the FAAH functional variant–BDNF association, suggests protein function alterations may have different biological consequences from expression changes, though this requires further investigation. Although colocalization was not supported across analyses, posterior probability patterns in several cases suggest limited power may have reduced detection of shared causal variants rather than true absence of biological relationship (31).

## 5. Strengths

A key strength is the systematic evaluation of the ECS across receptor genes and metabolic enzymes, providing broad assessment of ECS-related pathways in dementia. Using both functional missense variants and *cis*-eQTL instruments enabled investigation of complementary aspects of ECS biology — protein function and gene expression. Prioritising putatively functional variants (missense, synonymous, regulatory) enhances biological interpretability. Integrating clinical, biomarker, and neuroimaging outcome domains allowed assessment across different levels of disease biology, and the MR framework reduces confounding and reverse causation relative to observational studies (60). Colocalization analyses, though showing no shared variant evidence, enabled comprehensive evaluation.

## 6. Limitations

Limitations include limited genetic instruments for many genes, with several analyses relying on a single SNP, reducing power and precision. Null colocalization findings should be interpreted cautiously given assumptions underlying standard colocalization models (31).

Analyses were restricted to predominantly European ancestry populations, limiting generalisability. The primary findings were not independently replicated and require validation in independent datasets.

## 7. Conclusions

This study provides a systematic MR analysis of ECS-related genetic variation and dementia risk, and related neuroimaging and biomarker phenotypes. Although several associations survived correction, none was supported by colocalization. Nevertheless, biologically plausible signals were identified for *CNR2*, *FAAH*, *CNR1*, and *DAGLB*, suggesting selected ECS components may influence neurobiological pathways.

Overall, common ECS genetic variation is unlikely to have a major effect on dementia susceptibility but may contribute to specific neurodegeneration-related processes, including neuroinflammatory regulation, neurotrophic signalling, cerebrovascular function, and brain structural integrity. These findings highlight ECS biology’s complexity in dementia and support investigation in larger, well-powered studies integrating genetic, transcriptomic, proteomic, and functional data.

## Supporting information

STROBE MR

## Data Availability

Summary statistics from multiple genome-wide association studies were obtained from publicly available datasets. Details of all datasets included in the analyses are provided in Supplementary Tables 2 and 3. cis-eQTLs were obtained from the Genotype-Tissue Expression (GTEx) Project (Release V8), including brain, and adipose tissues. The GTEx Consortium atlas of genetic regulatory effects across human tissues was used as the primary source of eQTL data [GTEx Consortium, 2020]. Further information is available from the corresponding author upon reasonable request.

## List of Abbreviations

VasD: vascular dementia
AD: Alzheimer’s disease
ECS: endocannabinoid system
AEA: anandamide
2-AG: 2-arachidonoylglycerol
OEA: oleoylethanolamide
CB1: cannabinoid receptor type 1
CB2: cannabinoid receptor type 2
MR: Mendelian randomization
IVW: inverse-variance weighted
*cis*-: eQTL, *cis*-expression quantitative trait locus
IDP: imaging-derived phenotype
GWAS: genome-wide association study
FDR: false discovery rate
BH: Benjamini–Hochberg
APP: amyloid precursor protein
BDNF: brain-derived neurotrophic factor
NfL: neurofilament light chain
CSF: cerebrospinal fluid
GTEx: Genotype-Tissue Expression.

## Declarations

### Ethics approval and consent to participate

This study used publicly available summary-level genetic data and did not involve recruitment of participants or collection of new individual-level data. Ethical approval and informed consent were obtained in the original studies that generated the datasets used in this analysis.

### Consent for publication

Not applicable.

### Availability of data and materials

Summary statistics from multiple genome-wide association studies were obtained from publicly available datasets. Details of all datasets included in the analyses are provided in Supplementary Tables S3 and S4. *cis*-eQTLs were obtained from the Genotype-Tissue Expression (GTEx) Project (Release V8), including brain and adipose tissues. The GTEx Consortium atlas of genetic regulatory effects across human tissues was used as the primary source of eQTL data. Further information is available from the corresponding author upon reasonable request.

### Authors’ contributions

SJ developed the initial study structure and drafted the original manuscript. VK, BW, SB (Burgess), PP, LV and SB (Bhattacharyya) provided feedback, critically reviewed the manuscript and contributed to its revision. All authors read and approved the final manuscript.

## Acknowledgements

Not applicable.

## References

1. World Health Organization. Dementia. 2023.

2. Cummings J, Zhou Y, Lee G, Zhong K, Fonseca J, Cheng F. Alzheimer’s disease drug development pipeline: 2024. Alzheimer’s & Dementia: Translational Research & Clinical Interventions. 2024 Apr 24;10(2). doi:10.1002/trc2.12465

3. Livingston G, Huntley J, Liu KY, Costafreda SG, Selbæk G, Alladi S, et al. Dementia prevention, intervention, and care: 2024 report of the Lancet standing Commission. The Lancet. 2024 Aug;404(10452):572–628. doi:10.1016/S0140-6736(24)01296-0

4. Komedera M, Wojda U, Kiryk A. Cannabinoids in Alzheimer’s disease: animal–human evidence and clinical pharmacology challenges. Front Behav Neurosci. 2026 May 13;20. doi:10.3389/fnbeh.2026.1833021

5. Kanwal H, Sangineto M, Ciarnelli M, Castaldo P, Villani R, Romano A, et al. Potential Therapeutic Targets to Modulate the Endocannabinoid System in Alzheimer’s Disease. Int J Mol Sci. 2024 Apr 5;25(7):4050. doi:10.3390/ijms25074050

6. Mechoulam R, Parker LA. The Endocannabinoid System and the Brain. Annu Rev Psychol. 2013 Jan 3;64(1):21–47. doi:10.1146/annurev-psych-113011-143739

7. Di Marzo V. Endocannabinoids: synthesis and degradation. In. 2006. p. 1–24. doi:10.1007/112_0505

8. Lu HC, Mackie K. Review of the Endocannabinoid System. Biol Psychiatry Cogn Neurosci Neuroimaging. 2021 Jun;6(6):607–15. doi:10.1016/j.bpsc.2020.07.016

9. Benito C, Núñez E, Tolón RM, Carrier EJ, Rábano A, Hillard CJ, et al. Cannabinoid CB _2_ Receptors and Fatty Acid Amide Hydrolase Are Selectively Overexpressed in Neuritic Plaque-Associated Glia in Alzheimer’s Disease Brains. The Journal of Neuroscience. 2003 Dec 3;23(35):11136–41. doi:10.1523/JNEUROSCI.23-35-11136.2003

10. Ferreira PCL, Bellaver B, Povala G, Brum WS, Tissot C, Badji A, et al. Endocannabinoid System Biomarkers in Alzheimer’s Disease. Cannabis Cannabinoid Res. 2023 Feb;8(1):77–91. doi:10.1089/can.2022.0151 PubMed PMID: 36394442.

11. von Borcke N, Purwien AV, Steiert A, Hasecke H, Bouter Y. Reduced CB1 Cannabinoid Receptor Expression in Alzheimer’s Disease and Transgenic Mouse Models. AGING MEDICINE. 2026 Apr 24;9(2):168–84. doi:10.1002/agm2.70080

12. Piro JR, Benjamin DI, Duerr JM, Pi Y, Gonzales C, Wood KM, et al. A Dysregulated Endocannabinoid-Eicosanoid Network Supports Pathogenesis in a Mouse Model of Alzheimer’s Disease. Cell Rep. 2012 Jun;1(6):617–23. doi:10.1016/j.celrep.2012.05.001

13. Chen R, Zhang J, Wu Y, Wang D, Feng G, Tang YP, et al. Monoacylglycerol Lipase Is a Therapeutic Target for Alzheimer’s Disease. Cell Rep. 2012 Nov;2(5):1329–39. doi:10.1016/j.celrep.2012.09.030

14. Santos-García I, Rodríguez-Cueto C, Villegas P, Piscitelli F, Lauritano A, Shen CKJ, et al. Preclinical investigation in FAAH inhibition as a neuroprotective therapy for frontotemporal dementia using TDP-43 transgenic male mice. J Neuroinflammation. 2023 May 6;20(1):108. doi:10.1186/s12974-023-02792-z

15. Ferreira FF, Ribeiro FF, Rodrigues RS, Sebastião AM, Xapelli S. Brain-Derived Neurotrophic Factor (BDNF) Role in Cannabinoid-Mediated Neurogenesis. Front Cell Neurosci. 2018 Nov 28;12. doi:10.3389/fncel.2018.00441

16. Benyó Z, Ruisanchez É, Leszl-Ishiguro M, Sándor P, Pacher P. Endocannabinoids in cerebrovascular regulation. American Journal of Physiology-Heart and Circulatory Physiology. 2016 Apr 1;310(7):H785–801. doi:10.1152/ajpheart.00571.2015

17. Burgess S, Davey Smith G, Davies NM, Dudbridge F, Gill D, Glymour MM, et al. Guidelines for performing Mendelian randomization investigations: update for summer 2023. Wellcome Open Res. 2023 Aug 4;4:186. doi:10.12688/wellcomeopenres.15555.3

18. Karhunen V, Woolf B, Bhatnagar P, Gill D, Burgess S. Integrating genetic data with biological insight: A practical guide to cis-Mendelian randomization. The American Journal of Human Genetics. 2026 May;113(5):900–14. doi:10.1016/j.ajhg.2026.03.011

19. Cristino L, Becker T, Di Marzo V. Endocannabinoids and energy homeostasis: An update. BioFactors. 2014 Jul 8;40(4):389–97. doi:10.1002/biof.1168

20. Bartholdy A, Moseholm KF, Nielsen PY, Albrechtsen NJW, Gluud LL, Jensen MK. Long-term dementia risk in metabolic dysfunction-associated steatotic liver disease: a population-based study. Metab Brain Dis. 2026 Feb 2;41(1):25. doi:10.1007/s11011-026-01796-x

21. Skrivankova VW, Richmond RC, Woolf BAR, Yarmolinsky J, Davies NM, Swanson SA, et al. Strengthening the Reporting of Observational Studies in Epidemiology Using Mendelian Randomization. JAMA. 2021 Oct 26;326(16):1614. doi:10.1001/jama.2021.18236

22. Kunkle BW, Grenier-Boley B, Sims R, Bis JC, Damotte V, Naj AC, et al. Genetic meta-analysis of diagnosed Alzheimer’s disease identifies new risk loci and implicates Aβ, tau, immunity and lipid processing. Nat Genet. 2019 Mar 28;51(3):414–30. doi:10.1038/s41588-019-0358-2

23. Mega Vascular Cognitive Impairment and Dementia (MEGAVCID) consortium. A genome□wide association meta□ analysis of all□cause and vascular dementia. Alzheimer’s & Dementia. 2024 Sep 24;20(9):5973–95. doi:10.1002/alz.14115

24. Jansen IE, van der Lee SJ, Gomez-Fonseca D, de Rojas I, Dalmasso MC, Grenier-Boley B, et al. Genome-wide meta-analysis for Alzheimer’s disease cerebrospinal fluid biomarkers. Acta Neuropathol. 2022 Nov 6;144(5):821–42. doi:10.1007/s00401-022-02454-z

25. Sun BB, Chiou J, Traylor M, Benner C, Hsu YH, Richardson TG, et al. Plasma proteomic associations with genetics and health in the UK Biobank. Nature. 2023 Oct;622(7982):329–38. doi:10.1038/s41586-023-06592-6 PubMed PMID: 37794186.

26. Smith SM, Douaud G, Chen W, Hanayik T, Alfaro-Almagro F, Sharp K, et al. An expanded set of genome-wide association studies of brain imaging phenotypes in UK Biobank. Nat Neurosci. 2021 May 19;24(5):737–45. doi:10.1038/s41593-021-00826-4

27. Aguet F, Anand S, Ardlie KG, Gabriel S, Getz GA, Graubert A, et al. The GTEx Consortium atlas of genetic regulatory effects across human tissues. Science (1979). 2020 Sep 11;369(6509):1318–30. doi:10.1126/science.aaz1776

28. Chen Y, Lu T, Pettersson-Kymmer U, Stewart ID, Butler-Laporte G, Nakanishi T, et al. Genomic atlas of the plasma metabolome prioritizes metabolites implicated in human diseases. Nat Genet. 2023 Jan;55(1):44–53. doi:10.1038/s41588-022-01270-1 PubMed PMID: 36635386.

29. Verbanck M, Chen CY, Neale B, Do R. Detection of widespread horizontal pleiotropy in causal relationships inferred from Mendelian randomization between complex traits and diseases. Nat Genet. 2018;50(5):693–8. doi:10.1038/s41588-018-0099-7 PubMed PMID: 29686387.

30. Fisher RA. Statistical Methods for Research Workers. In. 1992. p. 66–70. doi:10.1007/978-1-4612-4380-9_6

31. Wallace C. Eliciting priors and relaxing the single causal variant assumption in colocalisation analyses. PLoS Genet. 2020 Apr;16(4):e1008720. doi:10.1371/journal.pgen.1008720 PubMed PMID: 32310995.

32. Zuber V, Grinberg NF, Gill D, Manipur I, Slob EAW, Patel A, et al. Combining evidence from Mendelian randomization and colocalization: Review and comparison of approaches. Am J Hum Genet. 2022 May 5;109(5):767–82. doi:10.1016/j.ajhg.2022.04.001 PubMed PMID: 35452592.

33. Benjamini Y, Hochberg Y. Controlling the False Discovery Rate: A Practical and Powerful Approach to Multiple Testing. J R Stat Soc Series B Stat Methodol. 1995 Jan 1;57(1):289–300. doi:10.1111/j.2517-6161.1995.tb02031.x

34. Vuic B, Milos T, Tudor L, Konjevod M, Nikolac Perkovic M, Jazvinscak Jembrek M, et al. Cannabinoid CB2 Receptors in Neurodegenerative Proteinopathies: New Insights and Therapeutic Potential. Biomedicines. 2022 Nov 22;10(12). doi:10.3390/biomedicines10123000 PubMed PMID: 36551756.

35. Komorowska-Müller JA, Schmöle AC. CB2 Receptor in Microglia: The Guardian of Self-Control. Int J Mol Sci. 2020 Dec 22;22(1):19. doi:10.3390/ijms22010019

36. Young AP, Denovan-Wright EM. The Dynamic Role of Microglia and the Endocannabinoid System in Neuroinflammation. Front Pharmacol. 2022 Feb 4;12. doi:10.3389/fphar.2021.806417

37. Bullock TA, Galpayage Dona KNU, Hale JF, Morales P, Jagerovic N, Andrews AM, et al. Activation of CB2R by synthetic CB2R agonist, PM289, improves brain endothelial barrier properties, decreases inflammatory response and enhances endothelial repair. NeuroImmune pharmacology and therapeutics. 2023 Dec;2(4):387–400. doi:10.1515/nipt-2023-0016 PubMed PMID: 38116176.

38. Koppel J, Vingtdeux V, Marambaud P, d’Abramo C, Jimenez H, Stauber M, et al. CB2 Receptor Deficiency Increases Amyloid Pathology and Alters Tau Processing in a Transgenic Mouse Model of Alzheimer’s Disease. Molecular Medicine. 2013 Jan 30;19(1):29–36. doi:10.2119/molmed.2013.00140

39. Schmöle AC, Lundt R, Ternes S, Albayram Ö, Ulas T, Schultze JL, et al. Cannabinoid receptor 2 deficiency results in reduced neuroinflammation in an Alzheimer’s disease mouse model. Neurobiol Aging. 2015 Feb;36(2):710–9. doi:10.1016/j.neurobiolaging.2014.09.019

40. Ruiz de Martín Esteban S, Benito-Cuesta I, Terradillos I, Martínez-Relimpio AM, Arnanz MA, Ruiz-Pérez G, et al. Cannabinoid CB2 Receptors Modulate Microglia Function and Amyloid Dynamics in a Mouse Model of Alzheimer’s Disease. Front Pharmacol. 2022 Apr 27;13. doi:10.3389/fphar.2022.841766

41. Ramirez SH, Haskó J, Skuba A, Fan S, Dykstra H, McCormick R, et al. Activation of Cannabinoid Receptor 2 Attenuates Leukocyte–Endothelial Cell Interactions and Blood–Brain Barrier Dysfunction under Inflammatory Conditions. The Journal of Neuroscience. 2012 Mar 21;32(12):4004–16. doi:10.1523/JNEUROSCI.4628-11.2012

42. Sipe JC, Chiang K, Gerber AL, Beutler E, Cravatt BF. A missense mutation in human fatty acid amide hydrolase associated with problem drug use. Proceedings of the National Academy of Sciences. 2002 Jun 11;99(12):8394–9. doi:10.1073/pnas.082235799

43. Milton NGN. Anandamide and noladin ether prevent neurotoxicity of the human amyloid-β peptide. Neurosci Lett. 2002 Oct;332(2):127–30. doi:10.1016/S0304-3940(02)00936-9

44. Jia J, Ma L, Wu M, Zhang L, Zhang X, Zhai Q, et al. Anandamide Protects HT22 Cells Exposed to Hydrogen Peroxide by Inhibiting CB1 Receptor-Mediated Type 2 NADPH Oxidase. Oxid Med Cell Longev. 2014;2014:1–16. doi:10.1155/2014/893516

45. Blázquez C, Chiarlone A, Bellocchio L, Resel E, Pruunsild P, García-Rincón D, et al. The CB1 cannabinoid receptor signals striatal neuroprotection via a PI3K/Akt/mTORC1/BDNF pathway. Cell Death Differ. 2015 Oct 20;22(10):1618–29. doi:10.1038/cdd.2015.11

46. Alqahtani SM, Al-kuraishy HM, Al Gareeb AI, Albuhadily AK, Alexiou A, Papadakis M, et al. Unlocking Alzheimer’s Disease: The Role of BDNF Signaling in Neuropathology and Treatment. Neuromolecular Med. 2025 May 17;27(1):36. doi:10.1007/s12017-025-08857-x

47. Piancatelli D, Aureli A, Sebastiani P, Colanardi A, Del Beato T, Del Cane L, et al. Gene- and Gender-Related Decrease in Serum BDNF Levels in Alzheimer’s Disease. Int J Mol Sci. 2022 Nov 23;23(23):14599. doi:10.3390/ijms232314599

48. Pak K, Kantonen T, Pekkarinen L, Nuutila P, Nummenmaa L. Association of CNR1 gene and cannabinoid 1 receptor protein in the human brain. J Neurosci Res. 2023 Mar 28;101(3):327–37. doi:10.1002/jnr.25149

49. Herkenham M, Lynn A, Johnson M, Melvin L, de Costa B, Rice K. Characterization and localization of cannabinoid receptors in rat brain: a quantitative in vitro autoradiographic study. The Journal of Neuroscience. 1991 Feb 1;11(2):563–83. doi:10.1523/JNEUROSCI.11-02-00563.1991

50. Cheng C, Yang C, Jia C, Wang Q. The Role of Cerebellum in Alzheimer’s Disease: A Forgotten Research Corner. Journal of Alzheimer’s Disease. 2023 Aug 29;95(1):75–8. doi:10.3233/JAD-230381

51. Braak H, Braak E. Neuropathological stageing of Alzheimer-related changes. Acta Neuropathol. 1991 Sep;82(4):239–59. doi:10.1007/BF00308809

52. Yang C, Liu G, Chen X, Le W. Cerebellum in Alzheimer’s disease and other neurodegenerative diseases: an emerging research frontier. MedComm (Beijing). 2024 Jul;5(7):e638. doi:10.1002/mco2.638 PubMed PMID: 39006764.

53. Liu G, Yang C, Wang X, Chen X, Cai H, Le W. Cerebellum in neurodegenerative diseases: Advances, challenges, and prospects. iScience. 2024 Nov;27(11):111194. doi:10.1016/j.isci.2024.111194

54. Lu HC, Mackie K. An Introduction to the Endogenous Cannabinoid System. Biol Psychiatry. 2016 Apr;79(7):516–25. doi:10.1016/j.biopsych.2015.07.028

55. Stumm C, Hiebel C, Hanstein R, Purrio M, Nagel H, Conrad A, et al. Cannabinoid receptor 1 deficiency in a mouse model of Alzheimer’s disease leads to enhanced cognitive impairment despite of a reduction in amyloid deposition. Neurobiol Aging. 2013 Nov;34(11):2574–84. doi:10.1016/j.neurobiolaging.2013.05.027

56. Reisenberg M, Singh PK, Williams G, Doherty P. The diacylglycerol lipases: structure, regulation and roles in and beyond endocannabinoid signalling. Philosophical Transactions of the Royal Society B: Biological Sciences. 2012 Dec 5;367(1607):3264–75. doi:10.1098/rstb.2011.0387

57. Scipioni L, Ciaramellano F, Carnicelli V, Leuti A, Lizzi AR, De Dominicis N, et al. Microglial Endocannabinoid Signalling in AD. Cells. 2022 Apr 6;11(7). doi:10.3390/cells11071237 PubMed PMID: 35406803.

58. Chen C. Inhibiting degradation of 2-arachidonoylglycerol as a therapeutic strategy for neurodegenerative diseases. Pharmacol Ther. 2023 Apr;244:108394. doi:10.1016/j.pharmthera.2023.108394

59. Nomura DK, Morrison BE, Blankman JL, Long JZ, Kinsey SG, Marcondes MCG, et al. Endocannabinoid Hydrolysis Generates Brain Prostaglandins That Promote Neuroinflammation. Science (1979). 2011 Nov 11;334(6057):809–13. doi:10.1126/science.1209200

60. Burgess S, Thompson SG. Interpreting findings from Mendelian randomization using the MR-Egger method. Eur J Epidemiol. 2017 May 19;32(5):377–89. doi:10.1007/s10654-017-0255-x

