## Supplementary figures for "Endocannabinoid system genes and dementia: a systematic Mendelian randomization study"

### Supplementary Figures Index

### Supplementary Figure S1. Mendelian randomization estimates for *DAGLA* functional variants in relation to Alzheimer's disease.

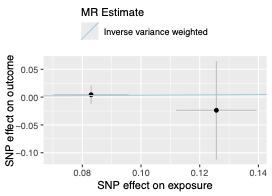

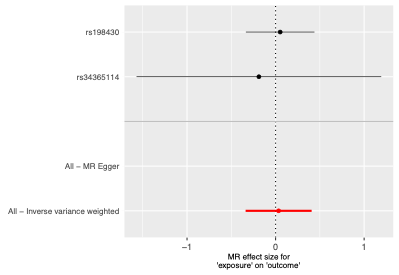

**Supplementary Figure S1.** Mendelian randomization analysis combining two *DAGLA* functional variants (rs198430 and rs34365114) using the inverse-variance weighted (IVW) method. The left panel shows a scatter plot of SNP effects on the exposure against SNP effects on Alzheimer's disease, with the IVW regression estimate. The right panel shows a forest plot of the individual SNP-specific estimates and the overall IVW estimate with 95% confidence intervals. IVW, inverse-variance weighted; MR, Mendelian randomization; SNP, single-nucleotide polymorphism.

### Supplementary Figure S2. Mendelian randomization estimates for *DAGLA* functional variants across neuroimaging-derived phenotypes from the UK Biobank.

#### A. Left nucleus accumbens volume (IDP 0023)

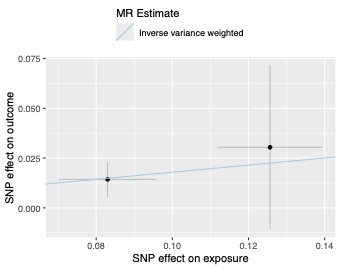

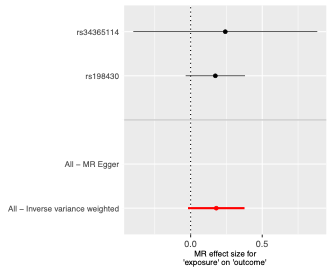

#### B. Left nucleus accumbens volume (IDP 0169)

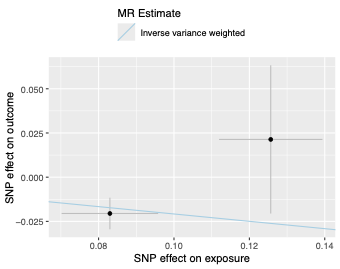

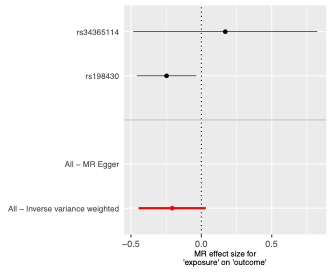

#### C. Left nucleus accumbens volume (IDP 0189)

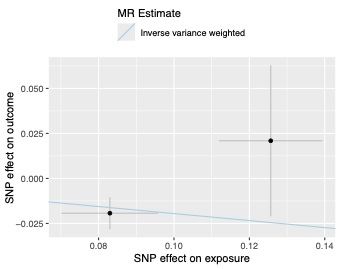

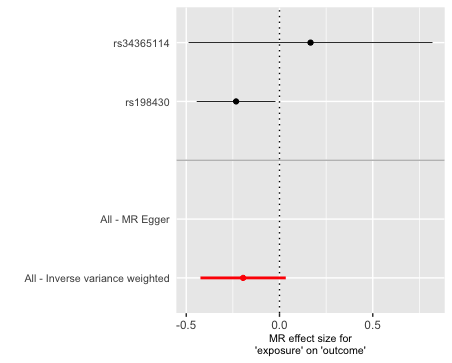

#### D. Left nucleus accumbens volume (IDP 0206)

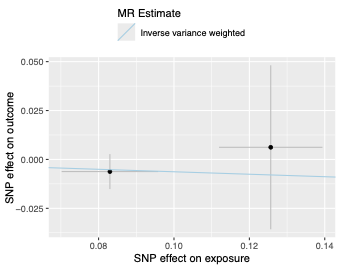

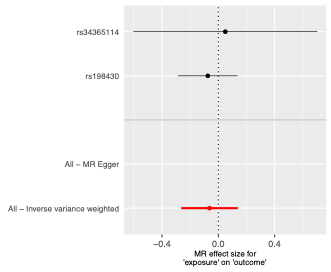

#### E. Left nucleus accumbens volume (IDP 1020)

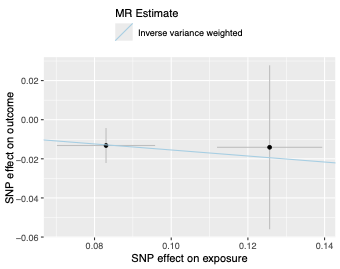

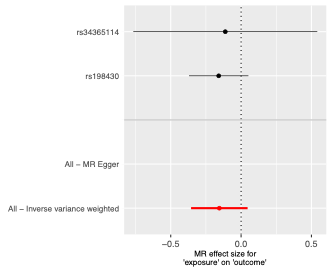

#### F. Right hippocampal volume (IDP 0286)

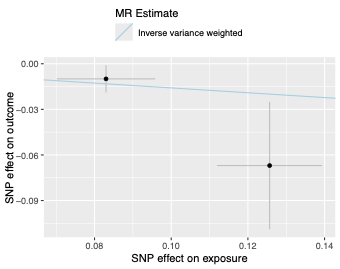

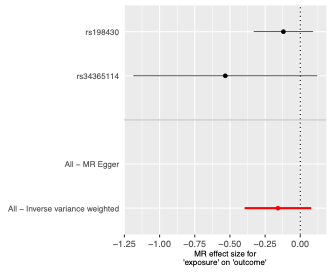

#### G. Brain segmentation volume (IDP 0165)

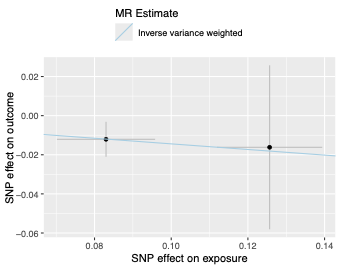

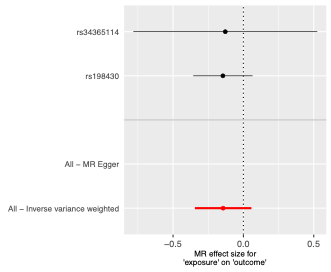

#### H. Right amygdala volume (IDP 0242)

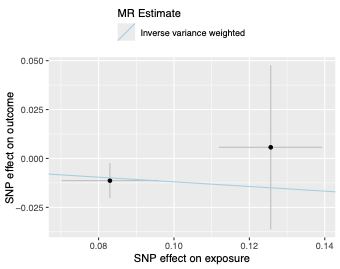

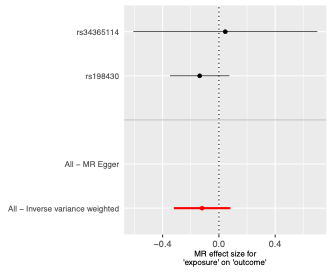

#### I. Left amygdala volume (IDP 0232)

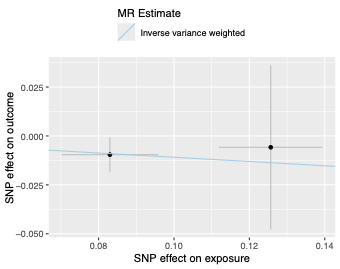

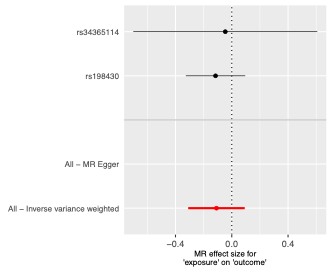

#### J. White matter hyperintensity volume (IDP 1437)

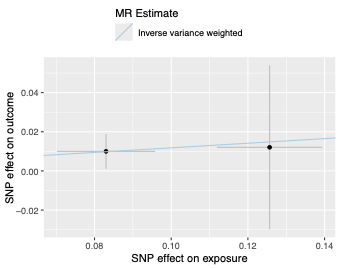

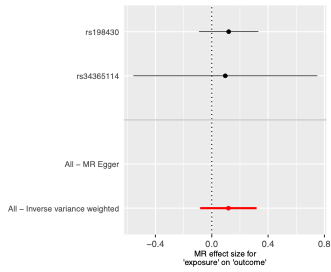

#### K. Whole brain volume (IDP 0009)

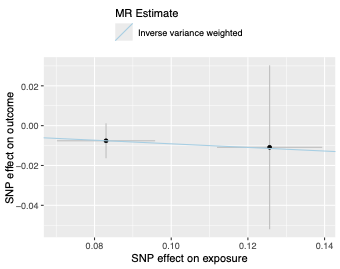

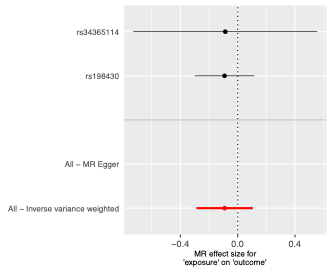

#### L. Left whole hippocampal volume (IDP 0264)

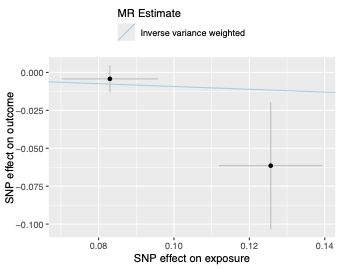

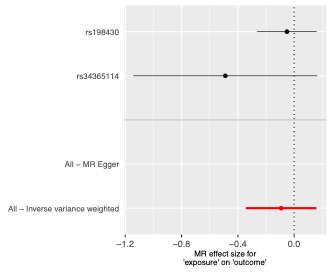

#### M. Right ventral striatum volume (IDP 0135)

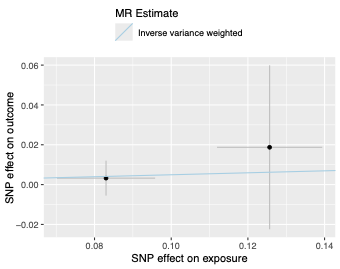

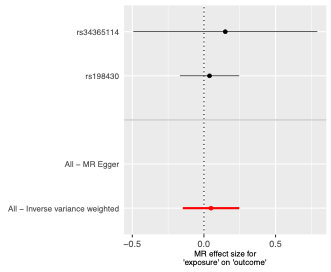

#### N. Left hippocampal volume (IDP 0019)

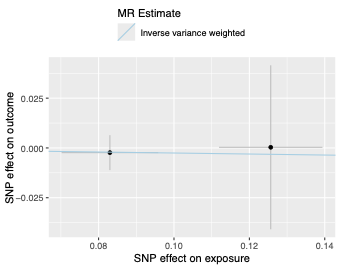

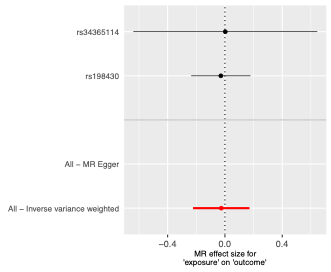

#### O. Left ventral striatum volume (IDP 0134)

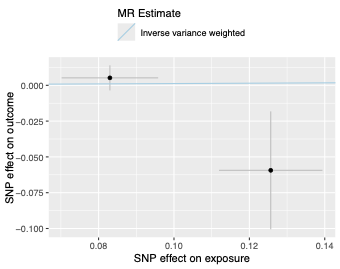

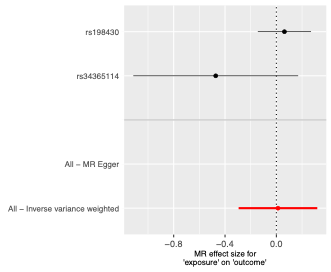

#### P. Right nucleus accumbens volume (IDP 0024)

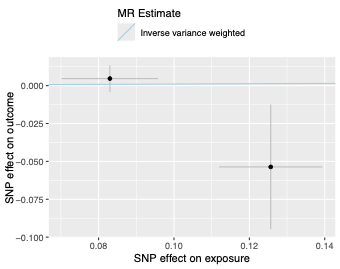

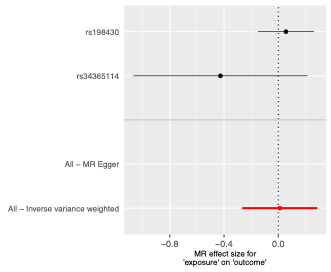

#### Q. Right hippocampal volume (IDP 0020)

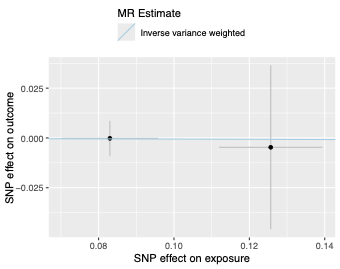

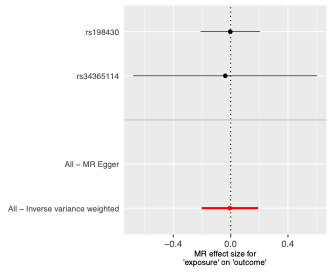

**Supplementary Figure S2.** Mendelian randomization analyses combining two *DAGLA* functional variants (rs198430 and rs34365114) using the inverse-variance weighted (IVW) method. Panels A–Q show analyses across selected UK Biobank neuroimaging-derived phenotypes. For each phenotype, the left panel shows a scatter plot of SNP effects on the exposure against SNP effects on the neuroimaging outcome, with the IVW regression estimate, and the right panel shows a forest plot of the individual SNP-specific estimates and the overall IVW estimate with 95% confidence intervals. IDP, imaging-derived phenotype; IVW, inverse-variance weighted; MR, Mendelian randomization; SNP, single-nucleotide polymorphism.

### Supplementary Figure S3. Mendelian randomization estimates for genetically predicted *CNR1* expression across tissues in relation to primary dementia outcomes.

#### A. *CNR1* expression in brain cerebellum in relation to Alzheimer's disease

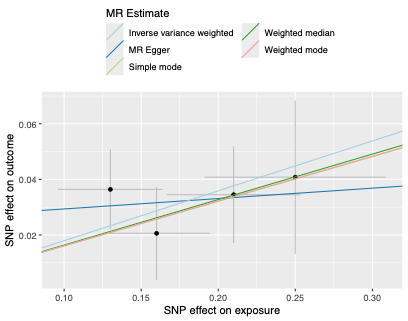

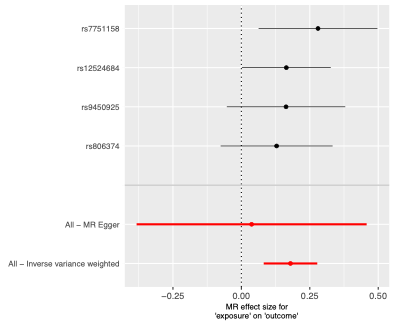

#### B. *CNR1* expression in brain cerebellar hemisphere in relation to Alzheimer's disease

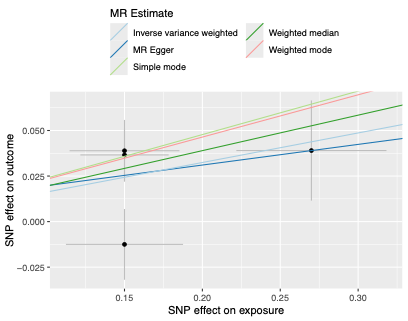

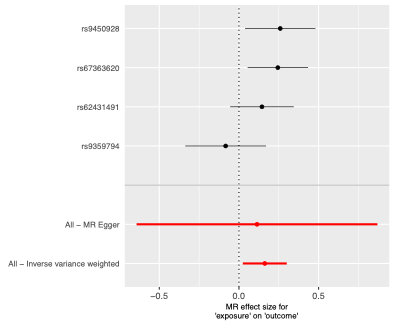

#### C. *CNR1* expression in brain basal ganglia in relation to Alzheimer's disease

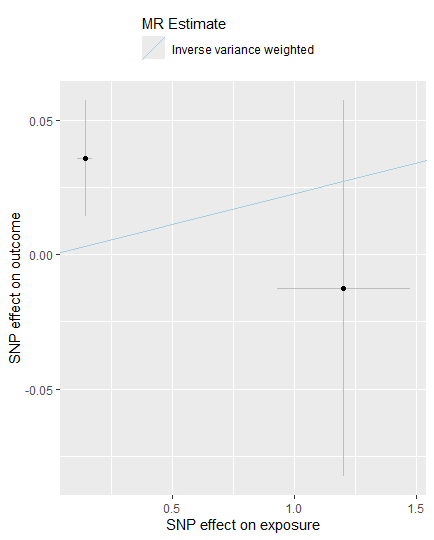

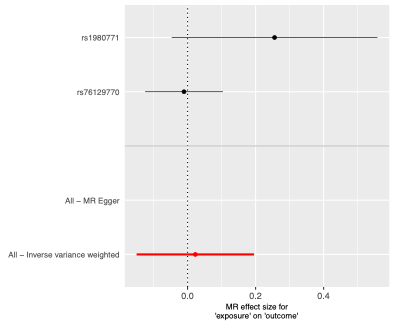

#### D. *CNR1* expression in subcutaneous adipose tissue in relation to Alzheimer's disease

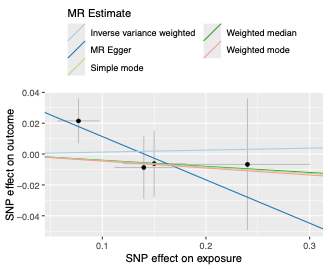

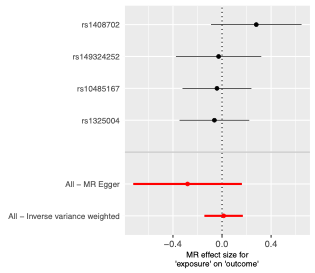

#### E. *CNR1* expression in subcutaneous adipose tissue in relation to vascular dementia

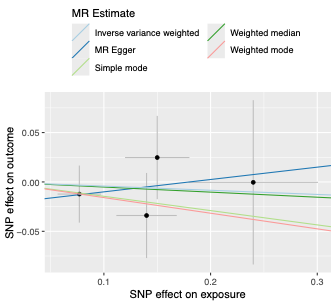

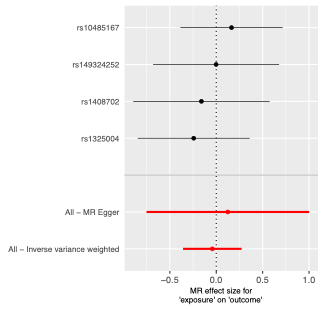

#### F. *CNR1* expression in brain cerebellum in relation to vascular dementia

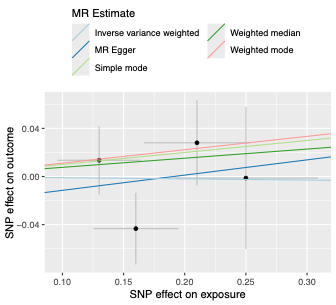

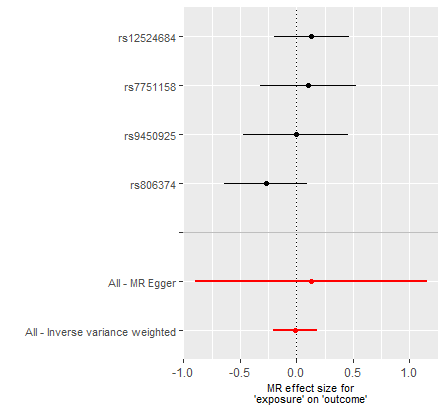

#### G. *CNR1* expression in brain cerebellar hemisphere in relation to vascular dementia

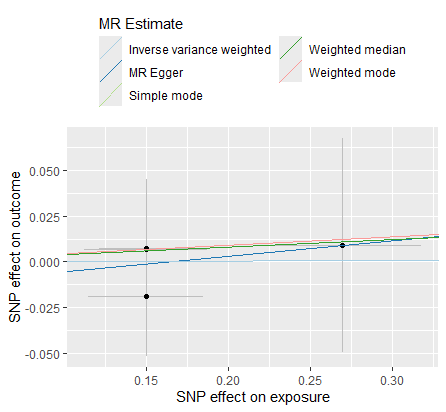

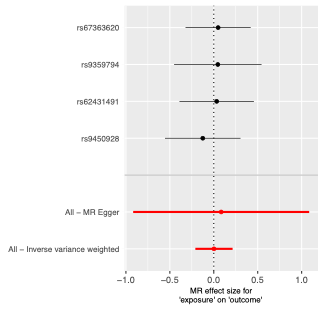

#### H. *CNR1* expression in brain basal ganglia in relation to all-cause dementia

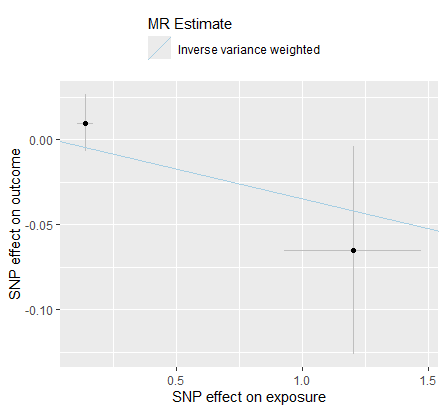

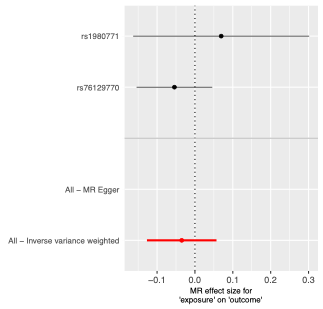

#### I. *CNR1* expression in subcutaneous adipose tissue in relation to all-cause dementia

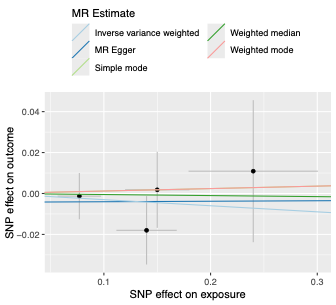

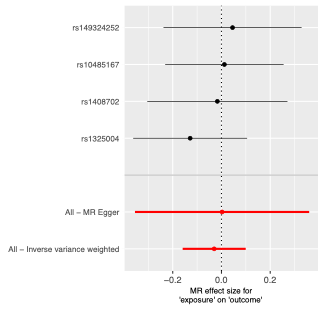

#### J. *CNR1* expression in brain cerebellar hemisphere in relation to all-cause dementia

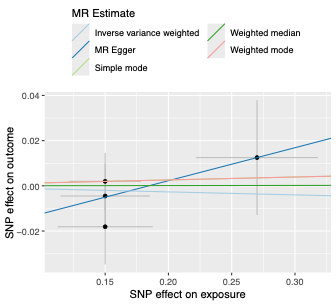

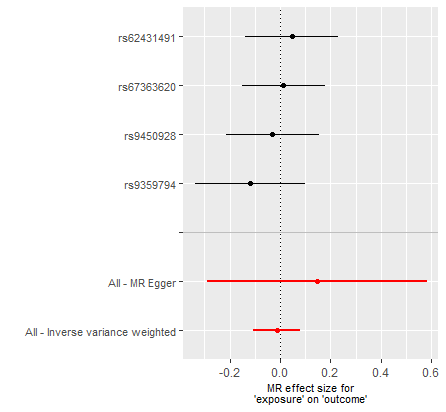

#### K. *CNR1* expression in brain cerebellum in relation to all-cause dementia

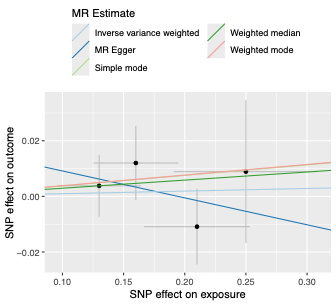

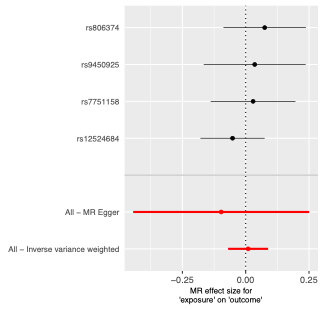

**Supplementary Figure S3.** Mendelian randomization analyses of genetically predicted *CNR1* expression across brain and adipose tissues in relation to Alzheimer's disease, vascular dementia, and all-cause dementia. Panels A–K show analyses across the evaluated tissue–outcome combinations. For each analysis, the left panel shows a scatter plot of SNP effects on *CNR1* expression against SNP effects on the corresponding dementia outcome, with regression lines for the applicable MR methods. The right panel shows a forest plot of the individual SNP-specific estimates and overall MR estimates with 95% confidence intervals. Where only two SNPs were available, estimates were obtained using the inverse-variance weighted (IVW) method only. IVW, inverse-variance weighted; MR, Mendelian randomization; SNP, single-nucleotide polymorphism.

### Supplementary Figure S4. Mendelian randomization estimates for genetically predicted *CNR1* expression in brain tissues in relation to secondary mechanistic outcomes.

#### A. *CNR1* expression in brain cerebellar hemisphere in relation to APP levels

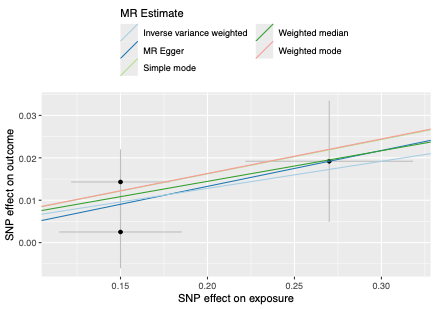

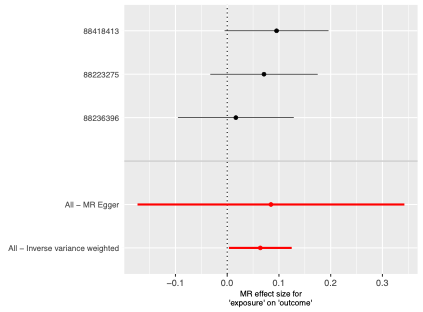

#### B. *CNR1* expression in brain cerebellum in relation to APP levels

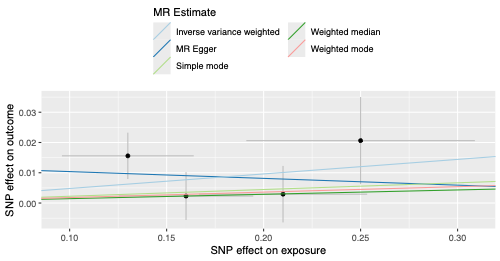

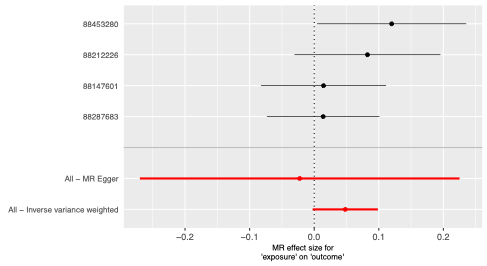

#### C. *CNR1* expression in brain cerebellar hemisphere in relation to BDNF levels

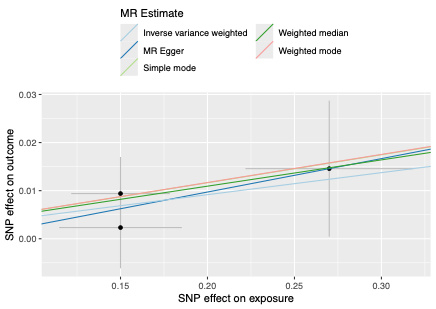

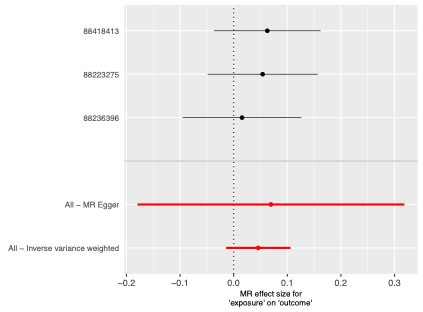

#### D. *CNR1* expression in brain cerebellum in relation to BDNF levels

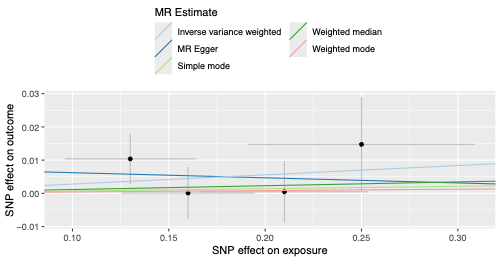

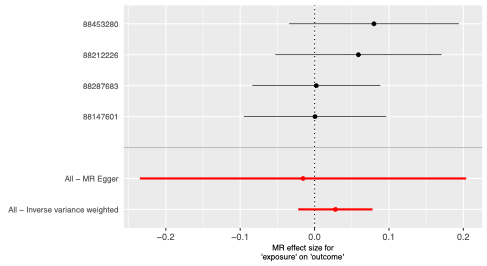

#### E. *CNR1* expression in brain cerebellar hemisphere in relation to GFAP levels

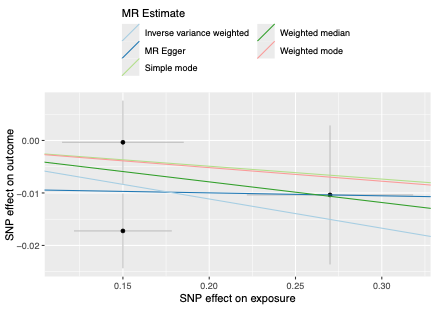

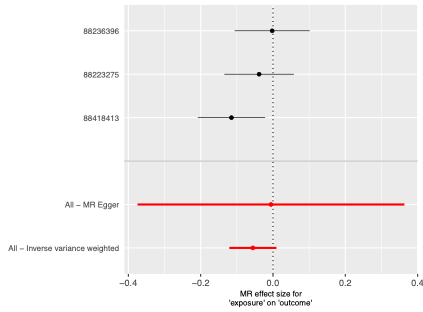

#### F. *CNR1* expression in brain cerebellum in relation to GFAP levels

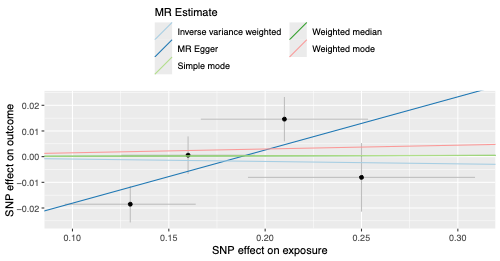

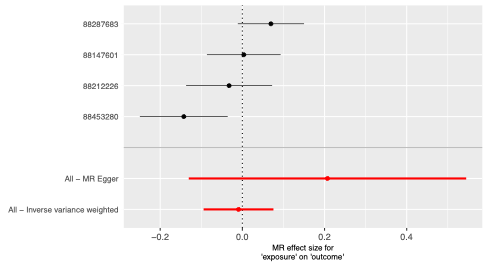

#### G. *CNR1* expression in brain cerebellar hemisphere in relation to MAPT levels

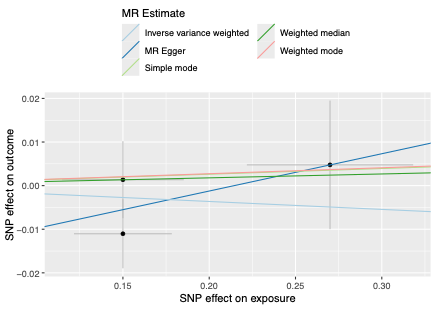

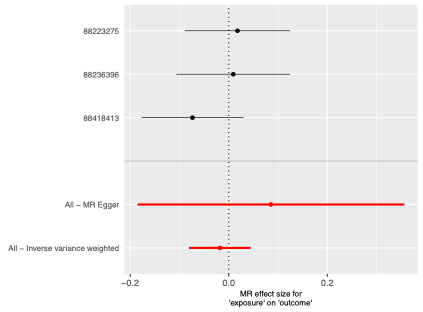

#### H. *CNR1* expression in brain cerebellum in relation to MAPT levels

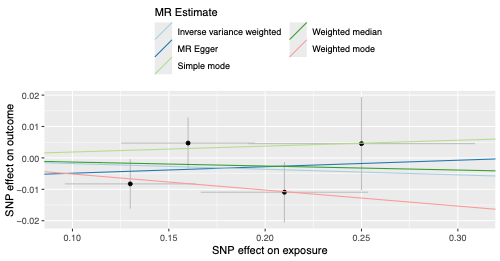

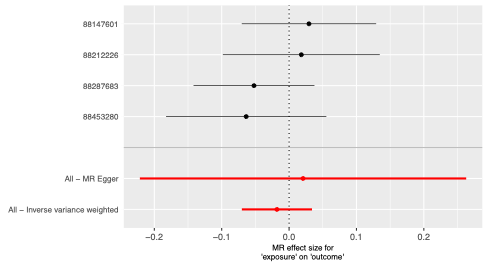

#### I. *CNR1* expression in brain cerebellar hemisphere in relation to NfL levels

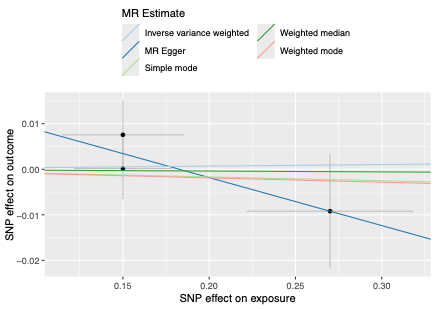

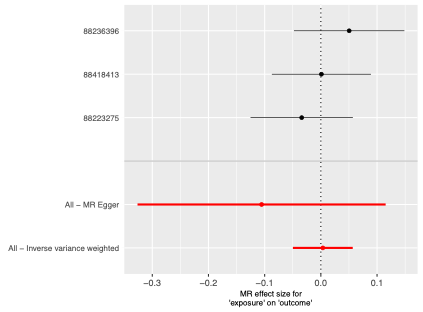

#### J. *CNR1* expression in brain cerebellum in relation to NfL levels

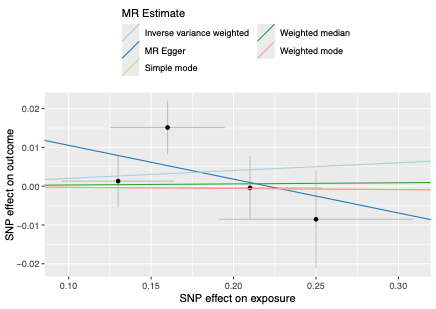

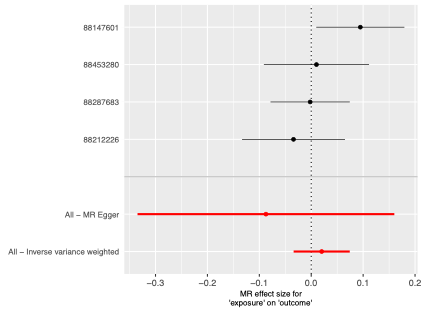

#### K. *CNR1* expression in brain cerebellar hemisphere in relation to Aβ levels

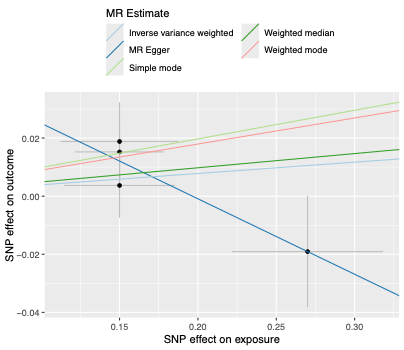

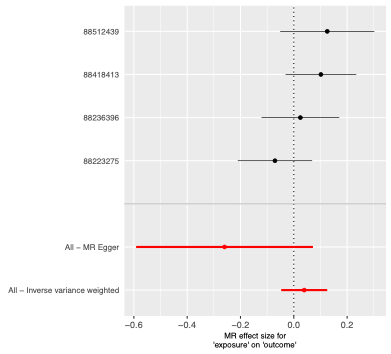

#### L. *CNR1* expression in brain cerebellum in relation to Aβ levels

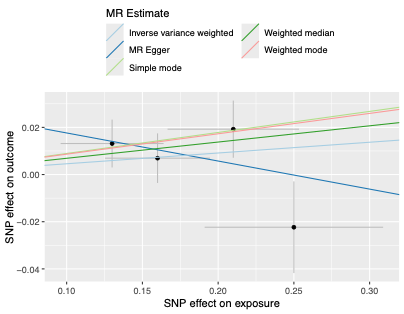

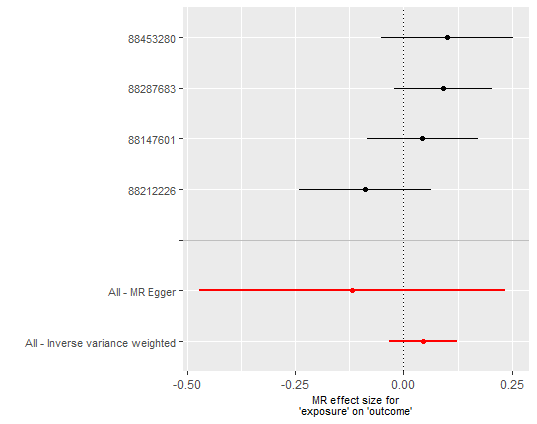

#### M. *CNR1* expression in brain cerebellar hemisphere in relation to tau levels

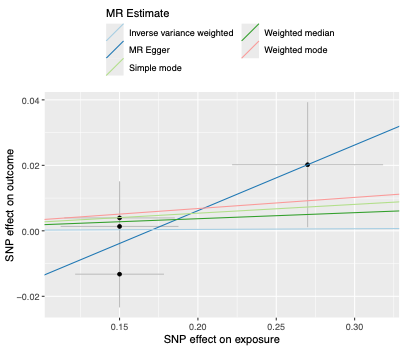

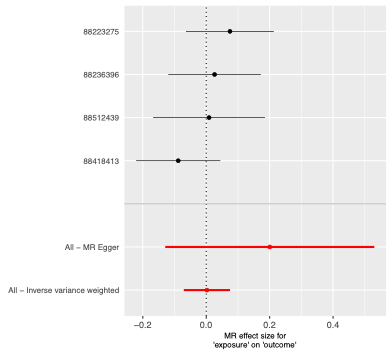

#### N. *CNR1* expression in brain cerebellum in relation to tau levels

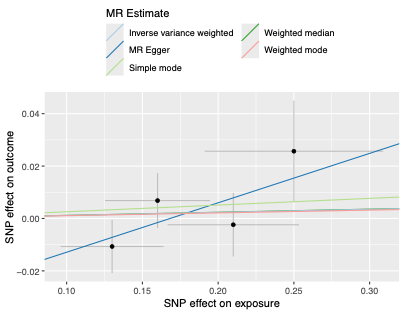

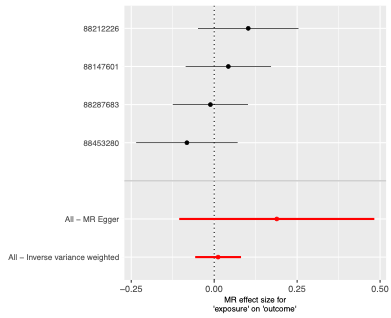

**Supplementary Figure S4.** Mendelian randomization analyses of genetically predicted CNR1 expression in the cerebellum and cerebellar hemisphere in relation to secondary mechanistic outcomes, including APP, BDNF, GFAP, MAPT, NfL, Aβ, and tau levels. Panels A–N show analyses across the evaluated tissue–outcome combinations. For each analysis, the left panel shows a scatter plot of SNP effects on *CNR1* expression against SNP effects on the corresponding mechanistic outcome, with regression lines for the applicable MR methods. The right panel shows a forest plot of the individual SNP-specific estimates and overall MR estimates with 95% confidence intervals. APP, amyloid precursor protein; Aβ, amyloid-beta; BDNF, brain-derived neurotrophic factor; GFAP, glial fibrillary acidic protein; MAPT, microtubule-associated protein tau; NfL, neurofilament light chain; IVW, inverse-variance weighted; MR, Mendelian randomization; SNP, single-nucleotide polymorphism.

### Supplementary Figure S5. Mendelian randomization estimates for genetically predicted *CNR1* expression in cerebellar tissues in relation to neuroimaging-derived phenotypes from the UK Biobank.

#### A. *CNR1* expression in brain cerebellum in relation to white matter hyperintensity volume (IDP 1437)

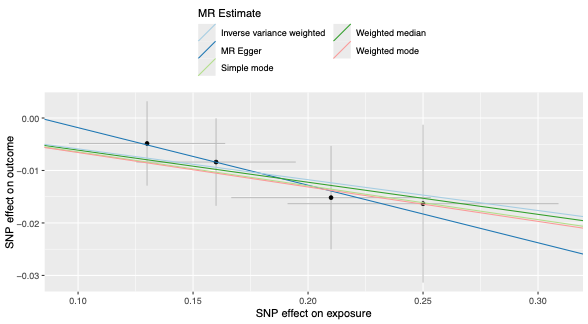

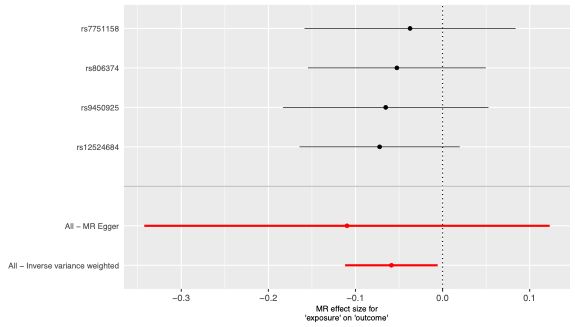

#### B. *CNR1* expression in brain cerebellum in relation to right whole hippocampal volume (IDP 0286)

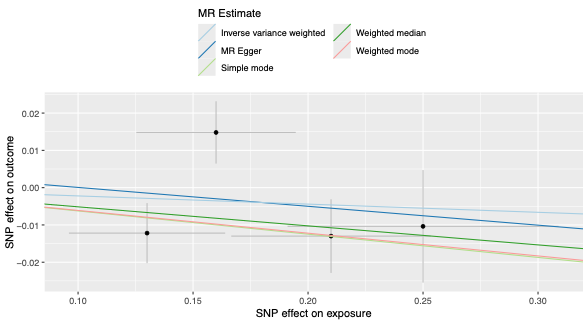

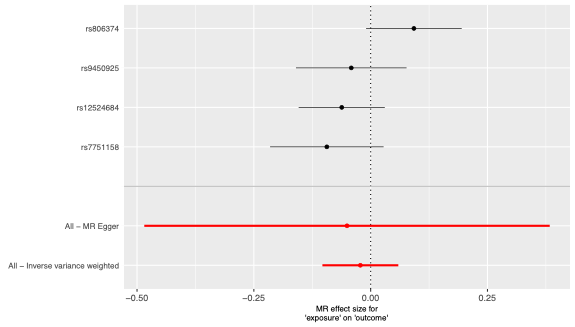

#### C. *CNR1* expression in brain cerebellum in relation to right ventral striatum grey matter volume (IDP 0135)

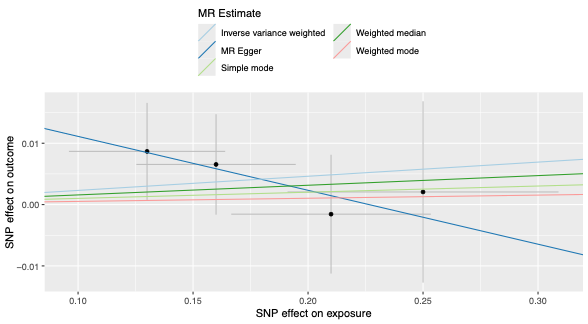

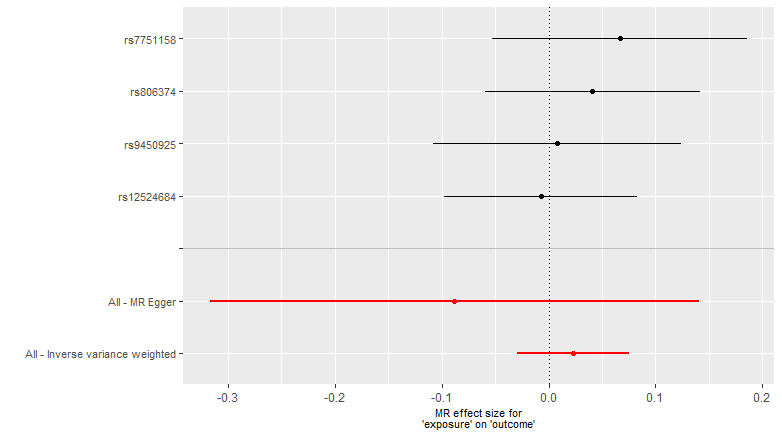

#### D. *CNR1* expression in brain cerebellum in relation to left nucleus accumbens volume (IDP 0023)

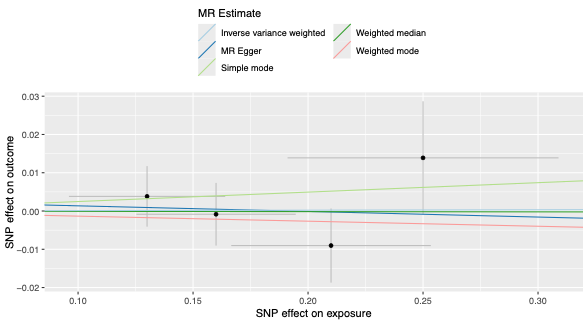

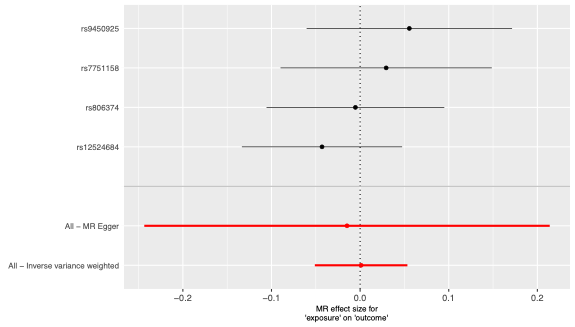

#### E. *CNR1* expression in brain cerebellum in relation to right hippocampal volume (IDP 0020)

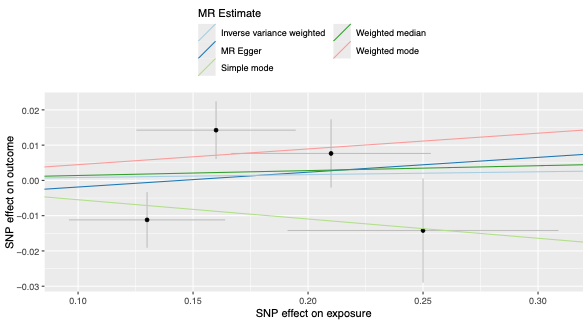

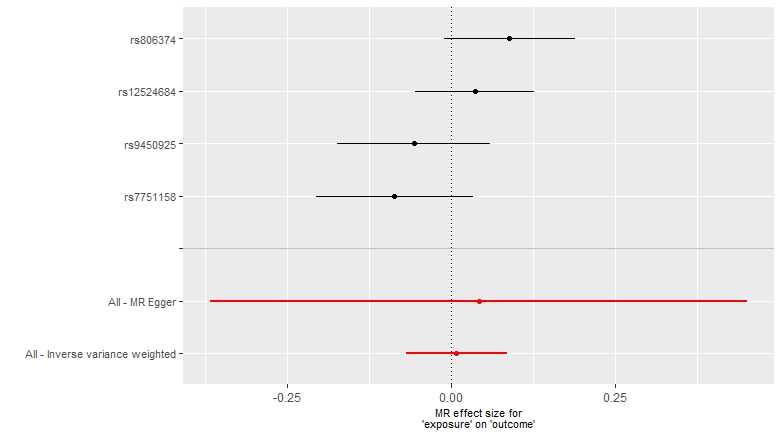

#### F. CNR1 expression in brain cerebellum in relation to left cortex volume (IDP 0189)

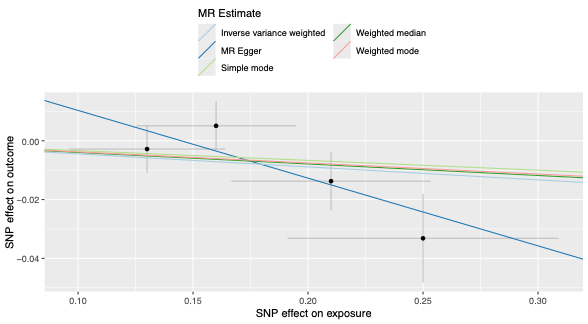

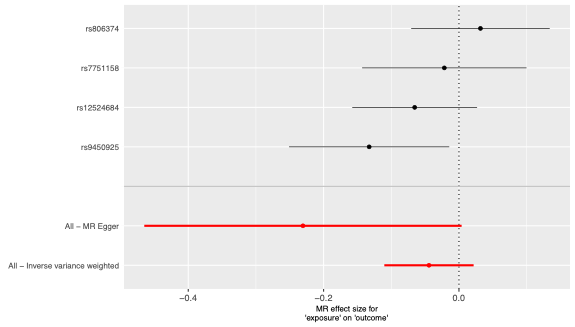

#### G. *CNR1* expression in brain cerebellum in relation to left ventral striatum grey matter volume (IDP 0134)

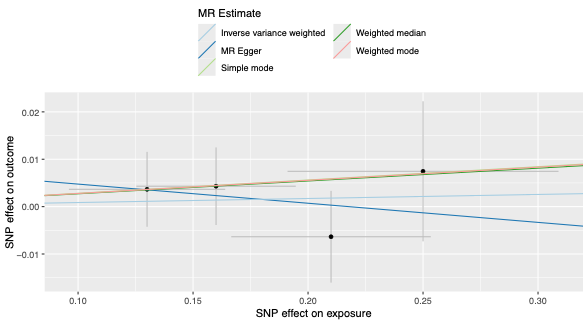

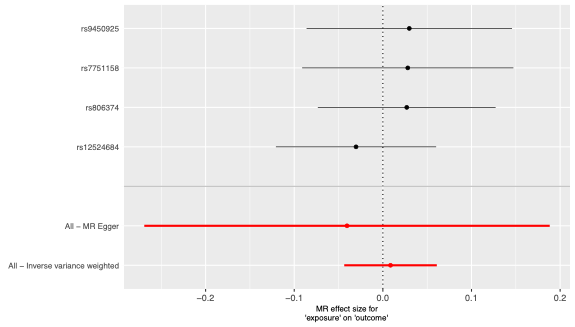

#### H. *CNR1* expression in brain cerebellum in relation to left hippocampal volume (IDP 0019)

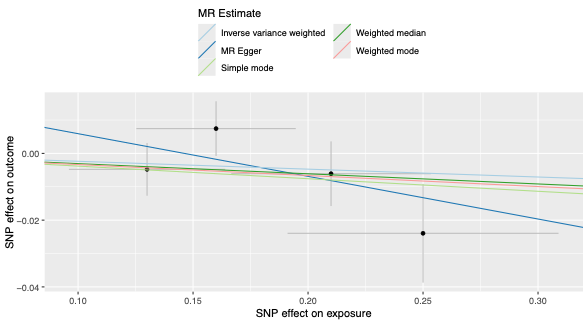

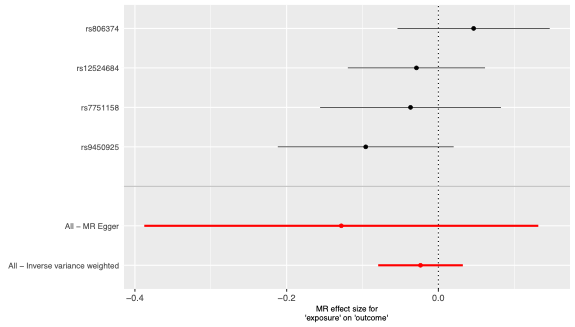

#### I. *CNR1* expression in brain cerebellum in relation to right whole amygdala volume

#### (IDP 0242)

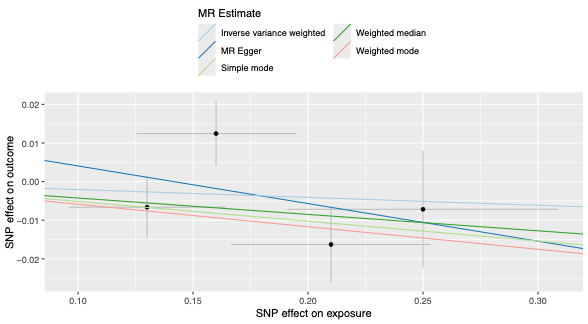

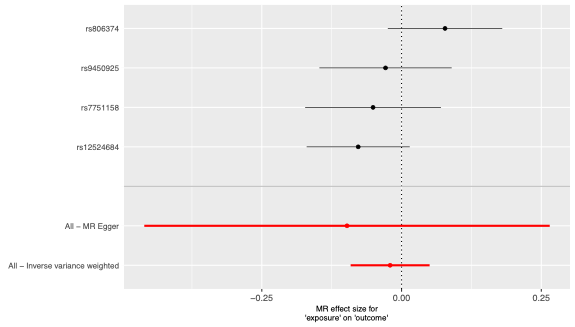

#### J. *CNR1* expression in brain cerebellum in relation to right cortex volume (IDP 0206)

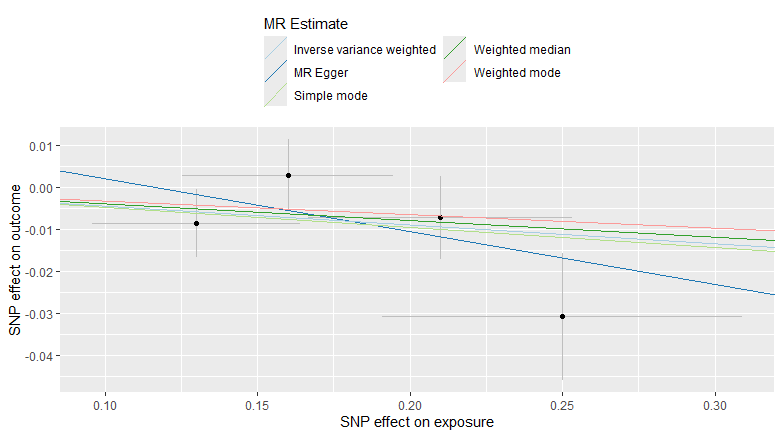

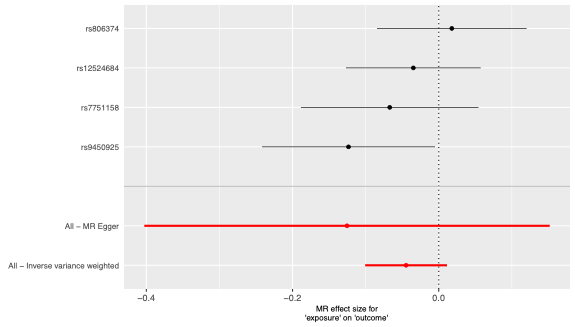

#### K. *CNR1* expression in brain cerebellum in relation to left whole hippocampal volume (IDP 0264)

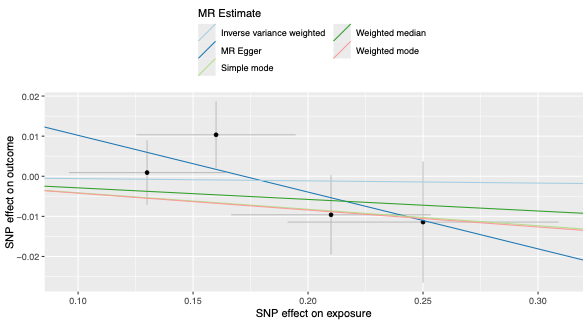

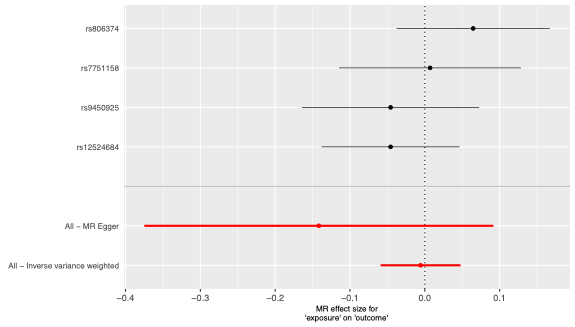

#### L. *CNR1* expression in brain cerebellum in relation to mean cortical thickness of the left hemisphere (IDP 1020)

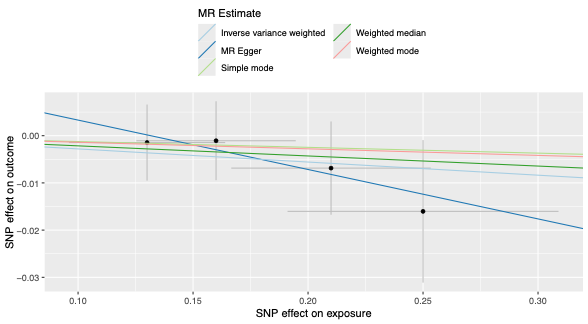

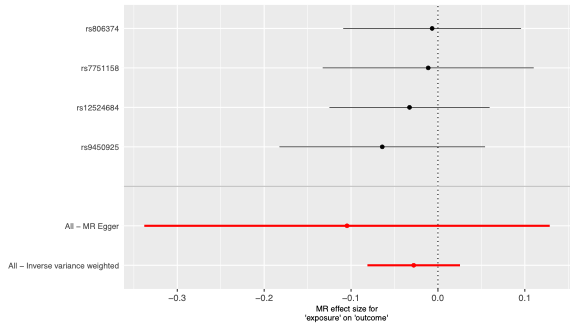

#### M. *CNR1* expression in brain cerebellum in relation to total grey matter volume (IDP 0169)

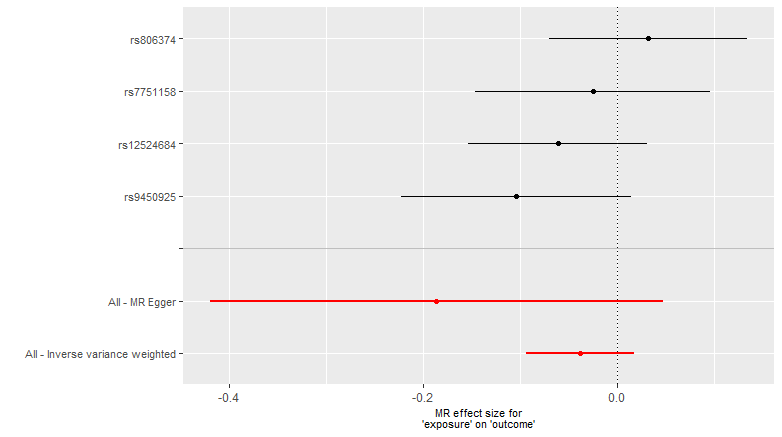

#### N. *CNR1* expression in brain cerebellum in relation to right nucleus accumbens volume (IDP 0024)

#### O. *CNR1* expression in brain cerebellum in relation to whole brain volume (IDP 0009)

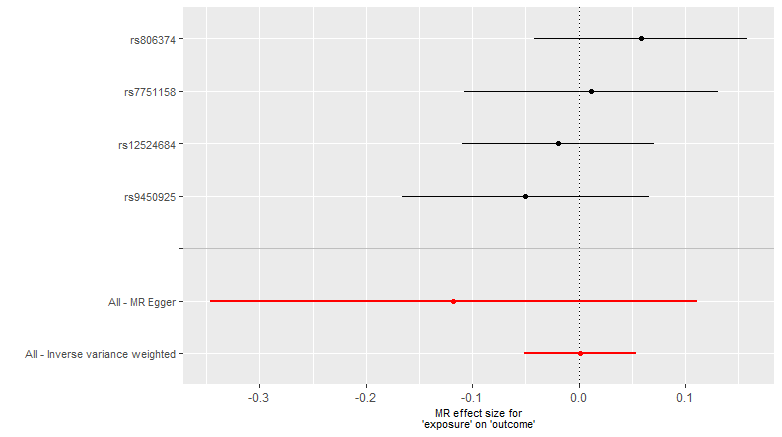

#### P. *CNR1* expression in brain cerebellum in relation to left whole amygdala volume (IDP 0232)

#### Q. *CNR1* expression in brain cerebellum in relation to brain segmentation-derived volume (IDP 0165)

#### R. *CNR1* expression in brain cerebellar hemisphere in relation to white matter hyperintensity volume (IDP 1437)

#### S. *CNR1* expression in brain cerebellar hemisphere in relation to right whole hippocampal volume (IDP 0286)

#### T. *CNR1* expression in brain cerebellar hemisphere in relation to right ventral striatum grey matter volume (IDP 0135)

#### U. *CNR1* expression in brain cerebellar hemisphere in relation to left nucleus accumbens volume (IDP 0023)

#### V. *CNR1* expression in brain cerebellar hemisphere in relation to right hippocampal volume (IDP 0020)

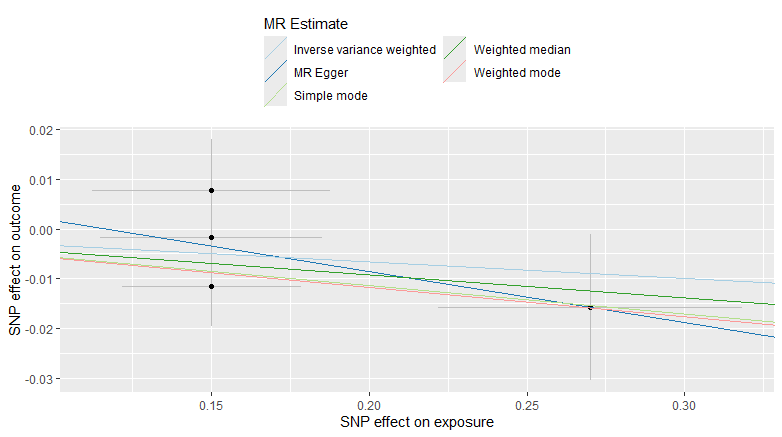

#### W. *CNR1* expression in brain cerebellar hemisphere in relation to left cortex volume (IDP 0189)

#### X. *CNR1* expression in brain cerebellar hemisphere in relation to left ventral striatum grey matter volume (IDP 0134)

#### Y. *CNR1* expression in brain cerebellar hemisphere in relation to left hippocampal volume (IDP 0019)

#### Z. *CNR1* expression in brain cerebellar hemisphere in relation to right whole amygdala volume (IDP 0242)

#### AA. *CNR1* expression in brain cerebellar hemisphere in relation to right cortex volume (IDP 0206)

#### AB. *CNR1* expression in brain cerebellar hemisphere in relation to left whole hippocampal volume (IDP 0264)

#### AC. *CNR1* expression in brain cerebellar hemisphere in relation to mean cortical thickness of the left hemisphere (IDP 1020)

#### AD. *CNR1* expression in brain cerebellar hemisphere in relation to total grey matter volume (IDP 0169)

#### AE. *CNR1* expression in brain cerebellar hemisphere in relation to right nucleus accumbens volume (IDP 0024)

#### AF. *CNR1* expression in brain cerebellar hemisphere in relation to whole brain volume (IDP 0009)

#### AG. *CNR1* expression in brain cerebellar hemisphere in relation to left whole amygdala volume (IDP 0232)

#### AH. *CNR1* expression in brain cerebellar hemisphere in relation to brain segmentation-derived volume (IDP 0165)

**Supplementary Figure S5.** Mendelian randomization analyses examining genetically predicted *CNR1* expression in brain cerebellum and brain cerebellar hemisphere in relation to neuroimaging-derived phenotypes from the UK Biobank. Panels A–Q show estimates for *CNR1* expression in brain cerebellum, and panels R–AH show estimates for *CNR1* expression in brain cerebellar hemisphere. For each analysis, the left panel shows a scatter plot of SNP effects on genetically predicted *CNR1* expression against SNP effects on the corresponding neuroimaging phenotype, with regression lines for the applied Mendelian randomization methods. The right panel shows a forest plot of the individual SNP-specific estimates together with the overall IVW and MR-Egger estimates and their 95% confidence intervals. IDP, imaging-derived phenotype; IVW, inverse-variance weighted; MR, Mendelian randomization; SNP, single-nucleotide polymorphism.

### Supplementary Figure S6. Mendelian randomization estimates for genetically predicted *DAGLB* expression across tissues in relation to primary dementia outcomes.

#### A. *DAGLB* expression in subcutaneous adipose tissue in relation to Alzheimer's disease

#### B. *DAGLB* expression in visceral adipose tissue in relation to Alzheimer's disease

#### C. *DAGLB* expression in brain cerebellar hemisphere in relation to Alzheimer's disease

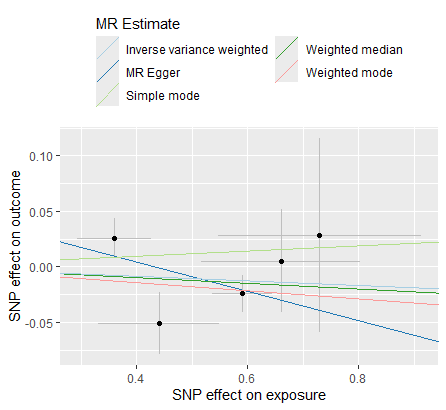

#### D. *DAGLB* expression in brain hippocampus in relation to Alzheimer's disease

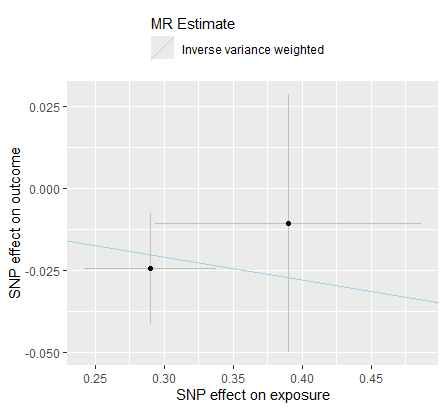

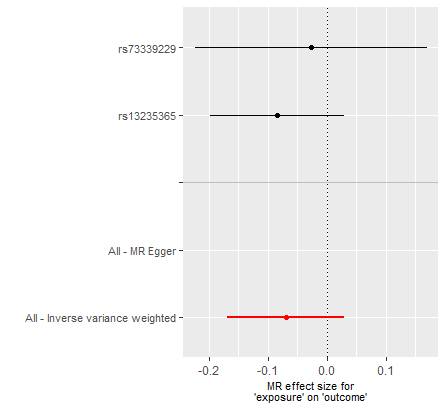

#### E. *DAGLB* expression in brain cerebellum in relation to Alzheimer's disease

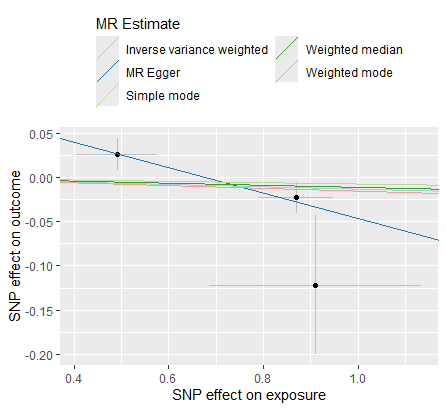

#### F. *DAGLB* expression in subcutaneous adipose tissue in relation to vascular dementia

#### G. *DAGLB* expression in visceral adipose tissue in relation to vascular dementia

#### H. *DAGLB* expression in brain cerebellar hemisphere in relation to vascular dementia

#### I. *DAGLB* expression in brain hippocampus in relation to vascular dementia

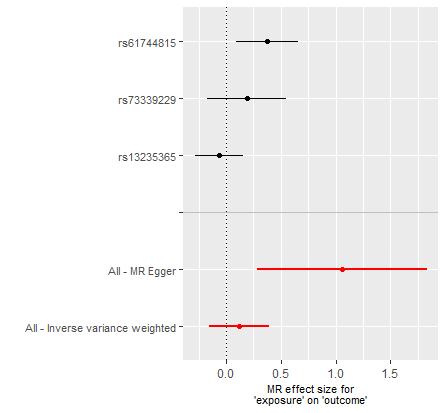

#### J. *DAGLB* expression in brain cerebellum in relation to vascular dementia

#### K. *DAGLB* expression in subcutaneous adipose tissue in relation to all-cause dementia

#### L. *DAGLB* expression in visceral adipose tissue in relation to all-cause dementia

#### M. *DAGLB* expression in brain cerebellar hemisphere in relation to all-cause dementia

#### N. *DAGLB* expression in brain hippocampus in relation to all-cause dementia

#### O. *DAGLB* expression in brain cerebellum in relation to all-cause dementia

**Supplementary Figure S6.** Mendelian randomization analyses examining genetically predicted *DAGLB* expression across subcutaneous adipose tissue, visceral adipose tissue, brain cerebellar hemisphere, brain hippocampus, and brain cerebellum in relation to Alzheimer’s disease, vascular dementia, and all-cause dementia. Panels A–E show estimates for Alzheimer’s disease, panels F–J for vascular dementia, and panels K–O for all-cause dementia. For each analysis, the left panel shows a scatter plot of SNP effects on genetically predicted *DAGLB* expression against SNP effects on the corresponding dementia outcome, with regression lines for the applied Mendelian randomization methods. The right panel shows a forest plot of the individual SNP-specific estimates together with the overall IVW and MR-Egger estimates and their 95% confidence intervals. IVW, inverse-variance weighted; MR, Mendelian randomization; SNP, single-nucleotide polymorphism.

### Supplementary Figure S7. Mendelian randomization estimates for genetically predicted *DAGLB* expression across tissues in relation to secondary mechanistic outcomes.

#### A. *DAGLB* expression in subcutaneous adipose tissue in relation to APP levels

#### B. *DAGLB* expression in visceral adipose tissue in relation to APP levels

#### C. *DAGLB* expression in brain cerebellar hemisphere in relation to APP levels

#### D. *DAGLB* expression in subcutaneous adipose tissue in relation to BDNF levels

#### E. *DAGLB* expression in visceral adipose tissue in relation to BDNF levels

#### F. *DAGLB* expression in brain cerebellar hemisphere in relation to BDNF levels

#### G. *DAGLB* expression in subcutaneous adipose tissue in relation to GFAP levels

#### H. *DAGLB* expression in visceral adipose tissue in relation to GFAP levels

#### I. *DAGLB* expression in brain cerebellar hemisphere in relation to GFAP levels

#### J. *DAGLB* expression in subcutaneous adipose tissue in relation to MAPT levels

#### K. *DAGLB* expression in visceral adipose tissue in relation to MAPT levels

#### L. *DAGLB* expression in brain cerebellar hemisphere in relation to MAPT levels

#### M. *DAGLB* expression in subcutaneous adipose tissue in relation to NfL levels

#### N. *DAGLB* expression in visceral adipose tissue in relation to NfL levels

#### O. *DAGLB* expression in brain cerebellar hemisphere in relation to NfL levels

#### P. *DAGLB* expression in subcutaneous adipose tissue in relation to Aβ levels

#### Q. *DAGLB* expression in visceral adipose tissue in relation to Aβ levels

#### R. *DAGLB* expression in brain cerebellar hemisphere in relation to Aβ levels

#### S. *DAGLB* expression in subcutaneous adipose tissue in relation to tau

#### T. *DAGLB* expression in visceral adipose tissue in relation to tau

#### U. *DAGLB* expression in brain cerebellar hemisphere in relation to tau

**Supplementary Figure S7.** Mendelian randomization analyses of genetically predicted *DAGLB* expression in subcutaneous adipose tissue, visceral adipose tissue, and brain cerebellar hemisphere in relation to secondary mechanistic outcomes, including APP, BDNF, GFAP, MAPT, NfL, Aβ, and tau levels. Panels A–U show analyses across the evaluated tissue–outcome combinations. For each analysis, the left panel shows a scatter plot of SNP effects on *DAGLB* expression against SNP effects on the corresponding mechanistic outcome, with regression lines for the applicable MR methods. The right panel shows a forest plot of the individual SNP-specific estimates and overall MR estimates with 95% confidence intervals. APP, amyloid precursor protein; Aβ, amyloid-beta; BDNF, brain-derived neurotrophic factor; GFAP, glial fibrillary acidic protein; MAPT, microtubule-associated protein tau; NfL, neurofilament light chain; IVW, inverse-variance weighted; MR, Mendelian randomization; SNP, single-nucleotide polymorphism.

### Supplementary Figure S8. Mendelian randomization estimates for genetically predicted *DAGLB* expression across tissues in relation to neuroimaging-derived phenotypes from the UK Biobank.

#### A. *DAGLB* expression in subcutaneous adipose tissue in relation to left whole amygdala volume (IDP 0232)

#### B. *DAGLB* expression in subcutaneous adipose tissue in relation to right whole amygdala volume (IDP 0242)

#### C. *DAGLB* expression in subcutaneous adipose tissue in relation to left ventral striatum grey matter volume (IDP 0134)

#### D. *DAGLB* expression in subcutaneous adipose tissue in relation to left whole hippocampal volume (IDP 0264)

#### E. *DAGLB* expression in subcutaneous adipose tissue in relation to brain segmentation-derived volume (IDP 0165)

#### F. *DAGLB* expression in subcutaneous adipose tissue in relation to right hippocampal volume (IDP 0020)

#### G. *DAGLB* expression in subcutaneous adipose tissue in relation to mean cortical thickness of the left hemisphere (IDP 1020)

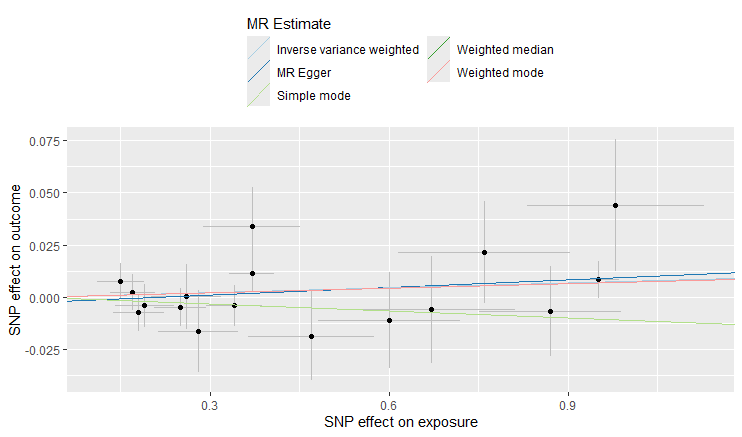

#### H. *DAGLB* expression in subcutaneous adipose tissue in relation to left hippocampal volume (IDP 0019)

#### I. *DAGLB* expression in subcutaneous adipose tissue in relation to right cortex volume (IDP 0206)

#### J. *DAGLB* expression in subcutaneous adipose tissue in relation to left nucleus accumbens volume (IDP 0023)

#### K. *DAGLB* expression in subcutaneous adipose tissue in relation to right whole hippocampal volume (IDP 0286)

#### L. *DAGLB* expression in subcutaneous adipose tissue in relation to left cortex volume (IDP 0189)

#### M. *DAGLB* expression in subcutaneous adipose tissue in relation to right ventral striatum grey matter volume (IDP 0135)

#### N. *DAGLB* expression in subcutaneous adipose tissue in relation to whole brain volume (IDP 0009)

#### O. *DAGLB* expression in subcutaneous adipose tissue in relation to white matter hyperintensity volume (IDP 1437)

#### P. *DAGLB* expression in subcutaneous adipose tissue in relation to total grey matter volume (IDP 0169)

#### Q. *DAGLB* expression in subcutaneous adipose tissue in relation to right nucleus accumbens volume (IDP 0024)

#### R. *DAGLB* expression in visceral adipose tissue in relation to left whole amygdala volume (IDP 0232)

#### S. *DAGLB* expression in visceral adipose tissue in relation to left ventral striatum grey matter volume (IDP 0134)

#### T. *DAGLB* expression in visceral adipose tissue in relation to right whole amygdala volume (IDP 0242)

#### U. *DAGLB* expression in visceral adipose tissue in relation to left hippocampal volume (IDP 0019)

#### V. *DAGLB* expression in visceral adipose tissue in relation to right cortex volume (IDP 0206)

#### W. *DAGLB* expression in visceral adipose tissue in relation to right ventral striatum grey matter volume (IDP 0135)

#### X. *DAGLB* expression in visceral adipose tissue in relation to total grey matter volume (IDP 0169)

#### Y. *DAGLB* expression in visceral adipose tissue in relation to brain segmentation-derived volume (IDP 0165)

#### Z. *DAGLB* expression in visceral adipose tissue in relation to left whole hippocampal volume (IDP 0264)

#### AA. *DAGLB* expression in visceral adipose tissue in relation to white matter hyperintensity volume (IDP 1437)

#### AB. *DAGLB* expression in visceral adipose tissue in relation to mean cortical thickness of the left hemisphere (IDP 1020)

#### AC. *DAGLB* expression in visceral adipose tissue in relation to right hippocampal volume (IDP 0020)

#### AD. *DAGLB* expression in visceral adipose tissue in relation to left nucleus accumbens volume (IDP 0023)

#### AE. *DAGLB* expression in visceral adipose tissue in relation to whole brain volume (IDP 0009)

#### AF. *DAGLB* expression in visceral adipose tissue in relation to right whole hippocampal volume (IDP 0286)

#### AG. *DAGLB* expression in visceral adipose tissue in relation to right nucleus accumbens volume (IDP 0024)

#### AH. *DAGLB* expression in visceral adipose tissue in relation to left cortex volume (IDP 0189)

#### AI. *DAGLB* expression in cerebellar hemisphere in relation to left whole amygdala volume (IDP 0232)

#### AJ. *DAGLB* expression in cerebellar hemisphere in relation to left ventral striatum grey matter volume (IDP 0134)

#### AK. *DAGLB* expression in cerebellar hemisphere in relation to left hippocampal volume (IDP 0019)

#### AL. *DAGLB* expression in cerebellar hemisphere in relation to right hippocampal volume (IDP 0020)

#### AM. *DAGLB* expression in cerebellar hemisphere in relation to right whole amygdala volume (IDP 0242)

#### AN. *DAGLB* expression in cerebellar hemisphere in relation to brain segmentation-derived volume (IDP 0165)

#### AO. *DAGLB* expression in cerebellar hemisphere in relation to whole brain volume (IDP 0009)

#### AP. *DAGLB* expression in cerebellar hemisphere in relation to left whole hippocampal volume (IDP 0264)

#### AQ. *DAGLB* expression in cerebellar hemisphere in relation to left cortex volume (IDP 0189)

#### AR. *DAGLB* expression in cerebellar hemisphere in relation to right nucleus accumbens volume (IDP 0024)

#### AS. *DAGLB* expression in cerebellar hemisphere in relation to total grey matter volume (IDP 0169)

#### AT. *DAGLB* expression in cerebellar hemisphere in relation to mean cortical thickness of the left hemisphere (IDP 1020)

#### AU. *DAGLB* expression in cerebellar hemisphere in relation to right cortex volume (IDP 0206)

#### AV. *DAGLB* expression in cerebellar hemisphere in relation to white matter hyperintensity volume (IDP 1437)

#### AW. *DAGLB* expression in cerebellar hemisphere in relation to left nucleus accumbens volume (IDP 0023)

#### AX. *DAGLB* expression in cerebellar hemisphere in relation to right whole hippocampal volume (IDP 0286)

#### AY. *DAGLB* expression in cerebellar hemisphere in relation to right ventral striatum grey matter volume (IDP 0135)

**Supplementary Figure S8.** Mendelian randomization analyses examining genetically predicted *DAGLB* expression in subcutaneous adipose tissue, visceral adipose tissue, and brain cerebellar hemisphere in relation to neuroimaging-derived phenotypes from the UK Biobank. Panels A–Q show estimates for *DAGLB* expression in subcutaneous adipose tissue, panels R–AH show estimates for *DAGLB* expression in visceral adipose tissue, and panels AI–AY show estimates for *DAGLB* expression in brain cerebellar hemisphere. For each analysis, the left panel shows a scatter plot of SNP effects on genetically predicted *DAGLB* expression against SNP effects on the corresponding neuroimaging phenotype, with regression lines for the applied Mendelian randomization methods. The right panel shows a forest plot of the individual SNP-specific estimates together with the overall IVW and MR-Egger estimates and their 95% confidence intervals. IDP, imaging-derived phenotype; IVW, inverse-variance weighted; MR, Mendelian randomization; SNP, single-nucleotide polymorphism.

### Supplementary Figure S9. Mendelian randomization estimates for genetically predicted *DAGLA* expression across tissues in relation to primary dementia outcomes.

#### A. *DAGLA* expression in subcutaneous adipose tissue in relation to Alzheimer’s disease

#### B. *DAGLA* expression in nucleus accumbens in relation to Alzheimer’s disease

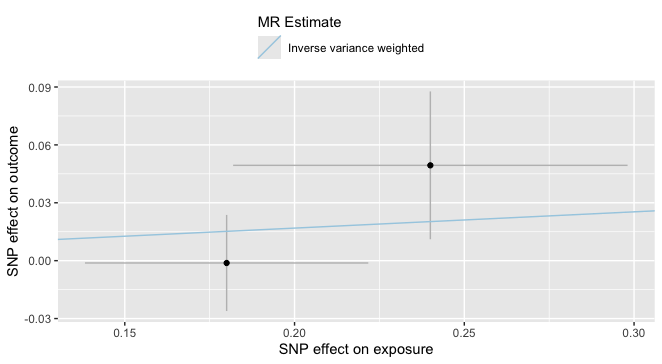

#### C. *DAGLA* expression in cerebellar hemisphere in relation to Alzheimer’s disease

#### D. *DAGLA* expression in cerebellum in relation to Alzheimer’s disease

#### E. *DAGLA* expression in caudate in relation to Alzheimer’s disease

#### F. *DAGLA* expression in cerebellar hemisphere in relation to vascular dementia

#### G. *DAGLA* expression in cerebellum in relation to vascular dementia

#### H. *DAGLA* expression in nucleus accumbens in relation to vascular dementia

#### I. *DAGLA* expression in caudate in relation to vascular dementia

#### J. *DAGLA* expression in cerebellar hemisphere in relation to all-cause dementia

#### K. *DAGLA* expression in nucleus accumbens in relation to all-cause dementia

#### L. *DAGLA* expression in cerebellum in relation to all-cause dementia

#### M. *DAGLA* expression in caudate in relation to all-cause dementia

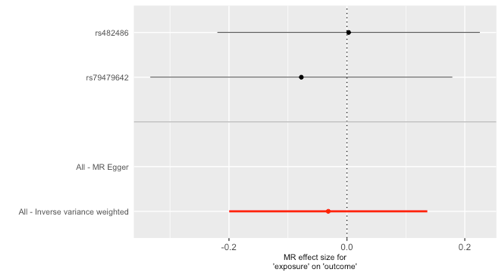

**Supplementary Figure S9.** Mendelian randomization analyses examining genetically predicted *DAGLA* expression across tissues in relation to primary dementia outcomes. Panels A–E show estimates for Alzheimer’s disease, panels F–I show estimates for vascular dementia, and panels J–M show estimates for all-cause dementia. For each analysis, the left panel shows a scatter plot of SNP effects on genetically predicted *DAGLA* expression against SNP effects on the corresponding dementia outcome, with regression lines for the applied Mendelian randomization methods. The right panel shows a forest plot of the individual SNP-specific estimates together with the overall IVW and MR-Egger estimates and their 95% confidence intervals. IVW, inverse-variance weighted; MR, Mendelian randomization; SNP, single-nucleotide polymorphism.

### Supplementary Figure S10. Mendelian randomization estimates for genetically predicted *FAAH* expression across tissues in relation to primary dementia outcomes.

#### A. *FAAH* expression in subcutaneous adipose tissue in relation to Alzheimer’s disease

#### B. *FAAH* expression in visceral adipose tissue in relation to Alzheimer’s disease

#### C. *FAAH* expression in cerebellar hemisphere in relation to Alzheimer’s disease

#### D. *FAAH* expression in cerebellum in relation to Alzheimer’s disease

#### E. *FAAH* expression in cortex in relation to Alzheimer’s disease

#### F. *FAAH* expression in hypothalamus in relation to Alzheimer’s disease

#### G. *FAAH* expression in putamen in relation to Alzheimer’s disease

#### H. *FAAH* expression in hippocampus in relation to Alzheimer’s disease

#### I. *FAAH* expression in caudate in relation to Alzheimer’s disease

#### J. *FAAH* expression in nucleus accumbens in relation to Alzheimer’s disease

#### K. *FAAH* expression in subcutaneous adipose tissue in relation to vascular dementia

#### L. *FAAH* expression in visceral adipose tissue in relation to vascular dementia

#### M. *FAAH* expression in cerebellar hemisphere in relation to vascular dementia

#### N. *FAAH* expression in cerebellum in relation to vascular dementia

#### O. *FAAH* expression in cortex in relation to vascular dementia

#### P. *FAAH* expression in hypothalamus in relation to vascular dementia

#### Q. *FAAH* expression in hippocampus in relation to vascular dementia

#### R. *FAAH* expression in nucleus accumbens in relation to vascular dementia

#### S. *FAAH* expression in putamen in relation to vascular dementia

#### T. *FAAH* expression in caudate in relation to vascular dementia

#### U. *FAAH* expression in subcutaneous adipose tissue in relation to all-cause dementia

#### V. *FAAH* expression in visceral adipose tissue in relation to all-cause dementia

#### W. *FAAH* expression in cerebellar hemisphere in relation to all-cause dementia

#### X. *FAAH* expression in cerebellum in relation to all-cause dementia

#### Y. *FAAH* expression in hypothalamus in relation to all-cause dementia

#### Z. *FAAH* expression in hippocampus in relation to all-cause dementia

#### AA. *FAAH* expression in cortex in relation to all-cause dementia

#### AB. *FAAH* expression in nucleus accumbens in relation to all-cause dementia

#### AC. *FAAH* expression in putamen in relation to all-cause dementia

**Supplementary Figure S10.** Mendelian randomization analyses examining genetically predicted *FAAH* expression across tissues in relation to primary dementia outcomes. Panels A–J show estimates for Alzheimer’s disease, panels K–T show estimates for vascular dementia, and panels U–AC show estimates for all-cause dementia. For each analysis, the left panel shows a scatter plot of SNP effects on genetically predicted *FAAH* expression against SNP effects on the corresponding dementia outcome, with regression lines for the applied Mendelian randomization methods. The right panel shows a forest plot of the individual SNP-specific estimates together with the overall IVW and MR-Egger estimates and their 95% confidence intervals. IVW, inverse-variance weighted; MR, Mendelian randomization; SNP, single-nucleotide polymorphism.

### Supplementary Figure S11. Mendelian randomization estimates for genetically predicted *NAPEPLD* expression across tissues in relation to primary dementia outcomes.

#### A. *NAPEPLD* expression in subcutaneous adipose tissue in relation to Alzheimer’s disease

#### B. *NAPEPLD* expression in visceral adipose tissue in relation to Alzheimer’s disease

#### C. *NAPEPLD* expression in cerebellar hemisphere in relation to Alzheimer’s disease

#### D. *NAPEPLD* expression in caudate in relation to Alzheimer’s disease

#### E. *NAPEPLD* expression in subcutaneous adipose tissue in relation to vascular dementia

#### F. *NAPEPLD* expression in visceral adipose tissue in relation to vascular dementia

#### G. *NAPEPLD* expression in cortex in relation to vascular dementia

#### H. *NAPEPLD* expression in spinal cord in relation to vascular dementia

#### I. *NAPEPLD* expression in subcutaneous adipose tissue in relation to all-cause dementia

#### J. *NAPEPLD* expression in visceral adipose tissue in relation to all-cause dementia

#### K. *NAPEPLD* expression in cerebellar hemisphere in relation to all-cause dementia

#### L. *NAPEPLD* expression in cortex in relation to all-cause dementia

#### M. *NAPEPLD* expression in caudate in relation to all-cause dementia

#### N. *NAPEPLD* expression in spinal cord in relation to all-cause dementia

**Supplementary Figure S11.** Mendelian randomization analyses examining genetically predicted *NAPEPLD* expression across tissues in relation to primary dementia outcomes. Panels A–D show estimates for Alzheimer’s disease, panels E–H show estimates for vascular dementia, and panels I–N show estimates for all-cause dementia. For each analysis, the left panel shows a scatter plot of SNP effects on genetically predicted *NAPEPLD* expression against SNP effects on the corresponding dementia outcome, with regression lines for the applied Mendelian randomization methods. The right panel shows a forest plot of the individual SNP-specific estimates together with the overall IVW and MR-Egger estimates and their 95% confidence intervals. IVW, inverse-variance weighted; MR, Mendelian randomization; SNP, single-nucleotide polymorphism.

### Supplementary Figure S12. Mendelian randomization estimates for genetically predicted *MGLL* expression across tissues in relation to primary dementia outcomes.

#### A. *MGLL* expression in visceral adipose tissue in relation to Alzheimer’s disease

#### B. *MGLL* expression in cerebellar hemisphere in relation to Alzheimer’s disease

#### C. *MGLL* expression in cerebellum in relation to Alzheimer’s disease

#### D. *MGLL* expression in nucleus accumbens in relation to Alzheimer’s disease

#### E. *MGLL* expression in putamen in relation to Alzheimer’s disease

#### F. *MGLL* expression in substantia nigra in relation to Alzheimer’s disease

#### G. *MGLL* expression in cerebellar hemisphere in relation to vascular dementia

#### H. *MGLL* expression in cerebellum in relation to vascular dementia

#### I. *MGLL* expression in nucleus accumbens in relation to vascular dementia

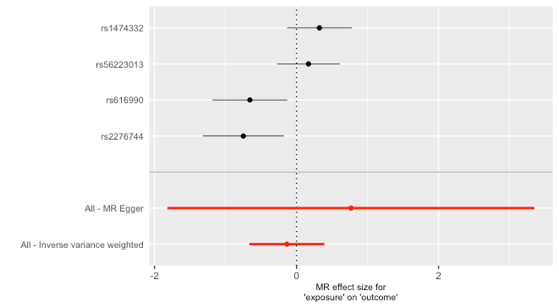

#### J. *MGLL* expression in putamen in relation to vascular dementia

#### K. *MGLL* expression in substantia nigra in relation to vascular dementia

#### L. *MGLL* expression in cerebellar hemisphere in relation to all-cause dementia

#### M. *MGLL* expression in cerebellum in relation to all-cause dementia

#### N. *MGLL* expression in nucleus accumbens in relation to all-cause dementia

#### O. *MGLL* expression in putamen in relation to all-cause dementia

#### P. *MGLL* expression in substantia nigra in relation to all-cause dementia

**Supplementary Figure S12.** Mendelian randomization analyses examining genetically predicted *MGLL* expression across tissues in relation to primary dementia outcomes. Panels A–F show estimates for Alzheimer’s disease, panels G–K show estimates for vascular dementia, and panels L–P show estimates for all-cause dementia. For each analysis, the left panel shows a scatter plot of SNP effects on genetically predicted *MGLL* expression against SNP effects on the corresponding dementia outcome, with regression lines for the applied Mendelian randomization methods. The right panel shows a forest plot of the individual SNP-specific estimates together with the overall IVW and MR-Egger estimates and their 95% confidence intervals. IVW, inverse-variance weighted; MR, Mendelian randomization; SNP, single-nucleotide polymorphism.

### Supplementary Figure S13. Colocalization analyses of cis-eQTL and GWAS signals for endocannabinoid system-related genes and selected outcomes.

#### A. Colocalization analysis of *CNR1* expression in brain cerebellum and Alzheimer’s disease

#### B. Colocalization analysis of *CNR1* expression in brain cerebellar hemisphere and Alzheimer’s disease.

#### C. Colocalization analysis of *CNR1* expression in brain cerebellar hemisphere and APP

#### D. Colocalization analysis of *DAGLB* expression in subcutaneous adipose tissue and all-cause dementia

#### E. Colocalization analysis of *DAGLB* expression in visceral adipose tissue and all-cause dementia

#### F. Colocalization analysis of *DAGLB* expression in subcutaneous adipose tissue and BDNF

#### G. Colocalization analysis of *DAGLB* expression in subcutaneous adipose tissue and NfL

#### H. Colocalization analysis of *DAGLB* expression in visceral adipose tissue and NfL

#### I. Colocalization analysis of *DAGLB* expression in subcutaneous adipose tissue and left amygdala volume

#### J. Colocalization analysis of *DAGLB* expression in visceral adipose tissue and left amygdala volume

#### K. Colocalization analysis of *DAGLB* expression in subcutaneous adipose tissue and left ventral striatum grey matter volume

#### L. Colocalization analysis of *DAGLB* expression in visceral adipose tissue and left ventral striatum grey matter volume

#### M. Colocalization analysis of *DAGLB* expression in cerebellar hemisphere and all-cause dementia

#### N. Colocalization analysis of *DAGLB* expression in cerebellar hemisphere and left amygdala volume

#### O. Colocalization analysis of *MGLL* expression in cortex and all-cause dementia

#### P. Colocalization analysis of *FAAH e*xpression in cortex and BDNF

1- *FAAH* cis- BDNF in caudate

#### Q. Colocalization analysis of *FAAH* expression in caudate and BDNF

#### R. Colocalization analysis of *FAAH* expression in cerebellum and BDNF

#### S. Colocalization analysis of *FAAH* expression in cerebellar hemisphere and BDNF

#### T. Colocalization analysis of *FAAH* expression in hippocampus and BDNF

#### U. Colocalization analysis of *FAAH* expression in hypothalamus and BDNF

#### V. Colocalization analysis of *FAAH* expression in nucleus accumbens and BDNF

#### W. Colocalization analysis of *FAAH* expression in putamen and BDNF

#### X. Colocalization analysis of *FAAH* expression in pituitary and BDNF

#### Y. Colocalization analysis of *FAAH* expression in subcutaneous adipose tissue and BDNF

#### Z. Colocalization analysis of *FAAH*-related OEA and BDNF

**Supplementary Figure S13.** Colocalization analyses of cis-eQTL and GWAS signals for endocannabinoid system-related genes and selected outcomes. Panels A–C show analyses for *CNR1* expression in relation to Alzheimer’s disease and APP, panels D–N show analyses for *DAGLB* expression in relation to all-cause dementia, BDNF, NfL, and selected neuroimaging-derived phenotypes, panel O shows the analysis for *MGLL* expression in relation to all-cause dementia, and panels P–Y show analyses for *FAAH* expression across tissues in relation to BDNF. Panel Z shows the colocalization analysis of *FAAH*-related OEA and BDNF. Each panel displays regional association signals for the corresponding genetic exposure and GWAS outcome across the genomic region used for colocalization analysis. APP, amyloid precursor protein; BDNF, brain-derived neurotrophic factor; cis-eQTL, cis-expression quantitative trait locus; GWAS, genome-wide association study; NfL, neurofilament light chain; OEA, oleoylethanolamide.
