## Supplementary methods for "Endocannabinoid system genes and dementia: a systematic Mendelian randomization study"

**M1. Selection of *cis*-eQTL instruments from GTEx.**

*C*is-expression quantitative trait loci (*cis*-eQTLs) were obtained from the Genotype-Tissue Expression (GTEx) project (version 8). In GTEx, *cis*-eQTL analyses were performed using FastQTL28, based on 838 donors with whole-genome sequencing data. Variants located within ±1 Mb of the transcription start site (TSS) and with MAF > 0.01 were considered. Analyses were adjusted for population structure using the first five principal components. Significant *c*is-eQTLs were extracted across tissues of interest, including brain and adipose tissues (Supplementary Table S19). To ensure independence of genetic instruments, linkage disequilibrium (LD) pruning was performed using the 1000 Genomes European reference panel (r² < 0.1), retaining the most strongly associated variant within each LD block.

**M2.** **Gene-specific methodological details.**

The analytical approach varied across functional variants according to the availability of exposure summary statistics, outcome data and suitable proxy variants. Gene-specific details are provided below.

***DAGLA***

Three putatively functional variants were identified for ***DAGLA*** (rs198430, rs34365114, and rs3741252). The synonymous variants rs198430 and rs34365114 were included in the Mendelian randomization analyses where exposure summary statistics were available. For vascular dementia and all-cause dementia, rs34365114 was unavailable in the outcome GWAS, and no suitable proxy variant (r² ≥ 0.8) could be identified; consequently, analyses for these outcomes were based on rs198430 alone. The missense variant rs3741252 was not included in the Mendelian randomization analyses because suitable exposure summary statistics were unavailable. Where individual variant association statistics were available, rs3741252 was evaluated using direct variant–outcome association analyses.

***DAGLB***

For *DAGLB* (rs1055430), MR analyses were performed for vascular dementia and all-cause dementia. For Alzheimer's disease, suitable MR analysis was not feasible because rs1055430 could not be retained as an instrumental variant following harmonisation. Consequently, the Alzheimer's disease estimate was obtained using a direct variant–outcome association analysis.

**M3. *FAAH*-specific colocalization analyses.**

The functional variant rs324420 was evaluated using two complementary colocalization approaches because it represents both a putatively functional missense variant and a *cis*-expression signal at the *FAAH* locus across multiple tissues. First, colocalization analyses were performed using *FAAH* *cis*-eQTLs across multiple tissues to assess whether genetically regulated *FAAH* expression shared a causal variant with the outcome. Second, because rs324420 is strongly associated with circulating oleoylethanolamide (OEA) levels and suitable GWAS summary statistics for anandamide (AEA) were unavailable, a GWAS of OEA was used as a proxy for FAAH enzymatic activity to assess whether genetically predicted FAAH activity shared a causal variant with the same outcome.
